# Whole person health across sociodemographic groups of adults in the United States

**DOI:** 10.64898/2026.10.13.26365413

**Authors:** Symielle A. Gaston, Christopher Payne, Chandra L. Jackson

## Abstract

Chronic disease accounts for most health care expenditures in the United States (US), highlighting the need for approaches that address health across interconnected spiritual, mental/emotional, behavioral, physical (including environmental), and social domains. Whole person health (WPH) is increasingly recognized as important for alleviating chronic disease burdens but has not been well-described across various sociodemographic groups of US adults. Using cross-sectional data from the 2025 National Health Interview Survey, we sought to describe the prevalence of WPH and its components among US adults overall and by age, sex, and race along with ethnicity. Participants responded to the nine-item Whole Person Health Index (WPHI), which we used to assess continuous scores (range: 9-45, where higher scores indicated improved health) and “optimal” (excellent/very good vs. good/fair/poor) overall and domain-specific WPH. Using survey procedures, we estimated medians (IQR) and prevalence (95% CI) and compared subgroups using multiple comparison correction. Among 23,884 NHIS participants aged 18 years and older (mean age (SE)=48 (0.2) years), representing approximately 261,620,640 US adults, the median WPHI score was 33 (28-37) and slightly over half (57% [56%-58%]) of adults were classified as having *“*optimal*”* overall WPH. Optimal social/family connections (72% [72%-73%]) were frequently reported; *“*optimal*”* sleep was infrequently reported (36% [35%-37%]). WPH and its components were generally highest among younger adults. Men and women differed in WPH domains but not in overall WPH. Non-Hispanic (NH) American Indian/Native Alaskan and NH-Black adults generally had lowest prevalence of optimal WPH and its components, which was often higher among Hispanic/Latino adults and highest among NH-Asian and White adults. These results showing variation in WPH in the US may inform future investigations of WPH, related predictors and health outcomes, and health disparities.

## INTRODUCTION

Despite decades of public health research focused on preventing, delaying, and reducing the burden of disease, health care expenditures on chronic disease in the United States (US) remain disproportionately high compared with those in other developed nations. Of the estimated $5.3 trillion spent on healthcare, chronic diseases account for 90% of expenditures.^1^ The impact of chronic disease extends beyond financial costs, contributing to disability, serious complications, and lost productivity.^1^ An estimated 76% of US adults have at least one chronic condition, and 51% of have two or more.^2^ These data indicate current approaches to addressing chronic disease have not been effective. The gap between healthcare costs and health outcomes underscores the need for novel approaches to help people reach salutogenesis or health.

The chronic disease epidemic may be partially explained by the traditional, disease-specific, siloed approach to healthcare. This approach often emphasizes pathogenesis and management of one condition at a time, rather than considering the broader set of interrelated factors that influence health and disease.^3–5^ There have recently been calls to move beyond siloed approaches to understanding health and disease toward integrative frameworks that employ systems approaches.^3–7^ These frameworks are designed to support comprehensive health and wellbeing as well as human and planetary flourishing.^3,5,6,8^ This shift requires a paradigm change: rather than focusing primarily on individual diseases, health should be understood across the continuum from salutogenesis to pathogenesis, with attention to the numerous interrelated causes that underlie this continuum.^3^ One such integrative framework is whole person health (WPH).

Although there is no standard definition of WPH, it can be described as health spanning across spiritual, mental/emotional, behavioral, physical, environmental, and social domains.^9,10^ In the WPH framework, health extends beyond the absence of a disease state but rather includes individuals’ total health, which is influenced by modifiable external and internal factors.^3–5,11^ As described in the Whole Well-being Model,^8^ WPH of an individual occurs within nested, interdependent layers. One larger layer, public health, includes shared environmental conditions, such as chemical stressors and health-related social needs, that influence systems and structures that affect WPH.^8^ Environmental conditions differ across sociodemographic groups, which in turn, likely contribute to disparities across many health outcomes,^12^ including in WPH. However, studies, to date, have not described WPH among US adults nor have they compared WPH across sociodemographic groups.

A prior study found that US adults consider lifestyle pillars such as sleep, physical activity, and nutrition important to their wellbeing.^13^ Each of these lifestyle pillars is a component of WPH and has been associated with multimorbidity.^10,14^ Another study described the prevalence of some but not all WPH-related activities (e.g., physical activity).^14^ However, these studies had several limitations. First, the available data did not include a comprehensive measure of WPH or important elements, such as religion and spirituality.^14^ Second, the assessments of engagement in WPH activities were not necessarily person-centered or directed since they did not evaluate perceived quality of WPH across domains, despite the premise that WPH reflects an individual’s values regarding health.^14^ Characterizing and describing WPH as reported by a nationally-representative sample of the US population is, therefore, important. Such a description provides an assessment of the current state of WPH among US adults and can contribute to the generation and testing of hypotheses about how WPH relates to the environment, health outcomes, and health disparities.

To address these literature gaps, we sought to describe the prevalence of WPH and its components overall and by age, sex, and race along with ethnicity among US adults in 2025, the most recent year in which WPH data are available in the National Health Interview Survey. Given the descriptive nature of this study, there was no *a priori* hypothesis. This descriptive study follows the previously published rigorous framework for descriptive epidemiology.^15^

## METHODS

### Data source

This analysis included cross-sectional data on sociodemographic characteristics and WPH from the 2025 National Health Interview Survey (NHIS)^16^ Details about the NHIS protocols and methodology, which were approved by the National Center for Health Statistics Ethics Review Board, are provided elsewhere.^17^ Briefly, NHIS is an annually administered survey used to capture health information about the civilian, non-institutionalized population of the United States (US). The NHIS employs a stratified, multistage probability sample to select individuals within US households for participation. All participants provided verbal consent to participate. In-person interviews were completed in respondents’ homes by trained field representatives from the US Census Bureau using computer-assisted personal interviewing. However, follow-up to complete interviews may have occurred via telephone. Due to closure of the federal government between October 1 and November 12, 2025, the 2025 NHIS sample included data collected over 11 months (January to September 30 and November 18 to December 7). For the 2025 NHIS adult sample, the weighted response rate was 41.3%.^17^ This analysis of secondary, de-identified data is not considered human subjects research; therefore, approval by the National Institutes of Health Institutional Review Board was waived. The authors’ analyses, interpretations, and conclusions are those of the authors and do not necessarily reflect the views of the National Center for Health Statistics, which is responsible only for the initial data.

### Study population

All 2025 NHIS adult participants aged 18 years and older (N=24,215) were eligible for the current study. However, exclusion criteria include missing data for the Whole Person Health Index^10^ or the assessed sociodemographic characteristics (age, sex, race, ethnicity). Therefore, n=331 were excluded due to missing data (Supplemental Figure 1), resulting in an analytic sample of 23,884 participants. Participants excluded for missing data were older but did not differ in racial and ethnic composition compared to participants in the analytic sample (Supplemental Table 1). Ten percent of excluded participants reported ‘don’t know’ or refused to provide their sex at birth.

### Sociodemographic characteristics

Self-reported sociodemographic characteristics considered stratification variables included age, sex, and race along with ethnicity (Table 1). We assessed age (years) continuously and using the following categories: 18-25; 26-44; 45-64; and ≥65 years. Participants reported sex (men, women) by responding to “Are you male or female?”. Lastly, we categorized the social construct of race and ethnicity based on participants’ self-identification, using broad Office of Management and Budget categories^18^ of Hispanic/Latino (any race and Hispanic/Latino ethnicity); non-Hispanic (NH)-American Indian/Native Alaskan (AIAN); NH-Asian; NH-Asian/Black African American (hereafter NH-Black); NH and multiple races or another racial group (e.g., Middle Eastern or North African, Native Hawaiian or Pacific Islander) than NH-AIAN, NH-Asian, NH-Black, or NH-White; and NH-White. While the NH-multiple races or another race group was analyzed, results are not interpreted due to high internal heterogeneity, which limits interpretability.

**Table 1.** Select sociodemographic characteristics, National Health Interview Survey, 2025 (N=23,884)

| Characteristics | Unweighted n | Prevalence (%) | Age-adjusted prevalence (%)<sup>a</sup> |
| --- | --- | --- | --- |
| Age (years), mean (standard error) | 23,884 | 48.2 (0.2) | - |
| Age group |  |  |  |
| 18-25 years | 1,634 | 13.4 (0.4) | - |
| 26-44 years | 6,788 | 32.7 (0.4) | - |
| 45-64 years | 7,125 | 30.8 (0.4) | - |
| ≥65 years | 8,337 | 23.1 (0.3) | - |
| Sex |  |  |  |
| Men | 11,071 | 48.7 (0.4) | 48.8 (0.4) |
| Women | 12,813 | 51.3 (0.4) | 51.2 (0.4) |
| Race and ethnicity |  |  |  |
| Hispanic/Latino | 3,364 | 18.6 (0.8) | 18.7 (0.8) |
| NH-American Indian/Alaskan Native | 158 | 0.6 (0.1) | 0.6 (0.1) |
| NH-Asian | 1,431 | 6.6 (0.3) | 6.6 (0.3) |
| NH-Black/African American | 2,680 | 11.9 (0.5) | 12.0 (0.5) |
| NH-Multiple or other race groups | 455 | 2.0 (0.1) | 2.1 (0.1) |
| NH-White | 15,796 | 60.3 (0.9) | 60.1 (0.9) |

Abbreviations: NH, non-Hispanic
All estimates are weighted for the survey's complex sampling design.
<sup>a</sup> Estimates except for age are age-standardized to the US 2020 population.

### Whole Person Health Index (WPHI)

The WPHI includes nine items that assess self-rated ability to find meaning and purpose in daily life; quality of life (QOL), focusing on what matters to respondents the most; general health; respondents’ health management, focusing on aspects of health that matter most to them; ability to manage stress; sleep; physical activity compared to people of the same age group; diet; and social and family connections.^10^ The WPHI has demonstrated strong internal consistency among US adults.^19^ Specific questions on WPHI were previously published and are provided in Table 2.^10^ NHIS participants responded to WPHI questions using a five-point Likert scale ranging from (1) excellent to (5) poor. Aligning with scoring guides,^10^ responses were reverse coded. Individual responses were summed to create a WPHI composite score ranging from 9 to 45, where higher scores indicated improving health. For each WPHI item, we assessed the individual score (range=1-5). In addition to the composite WPHI score, we also assessed “optimal” WPH by averaging the scores across the nine WPHI items, rounding it to the nearest integer, and dichotomizing that integer as ≥4 (corresponding to excellent/very good) vs. ≤3 (corresponding to good/fair/poor). We used this approach since there is no identified standard threshold of the WPHI that delineates “optimal” versus less than “optimal” WPH. Lastly, we considered “optimal” values (excellent/very good vs. good/fair/poor) of each WPHI item.

**Table 2.**
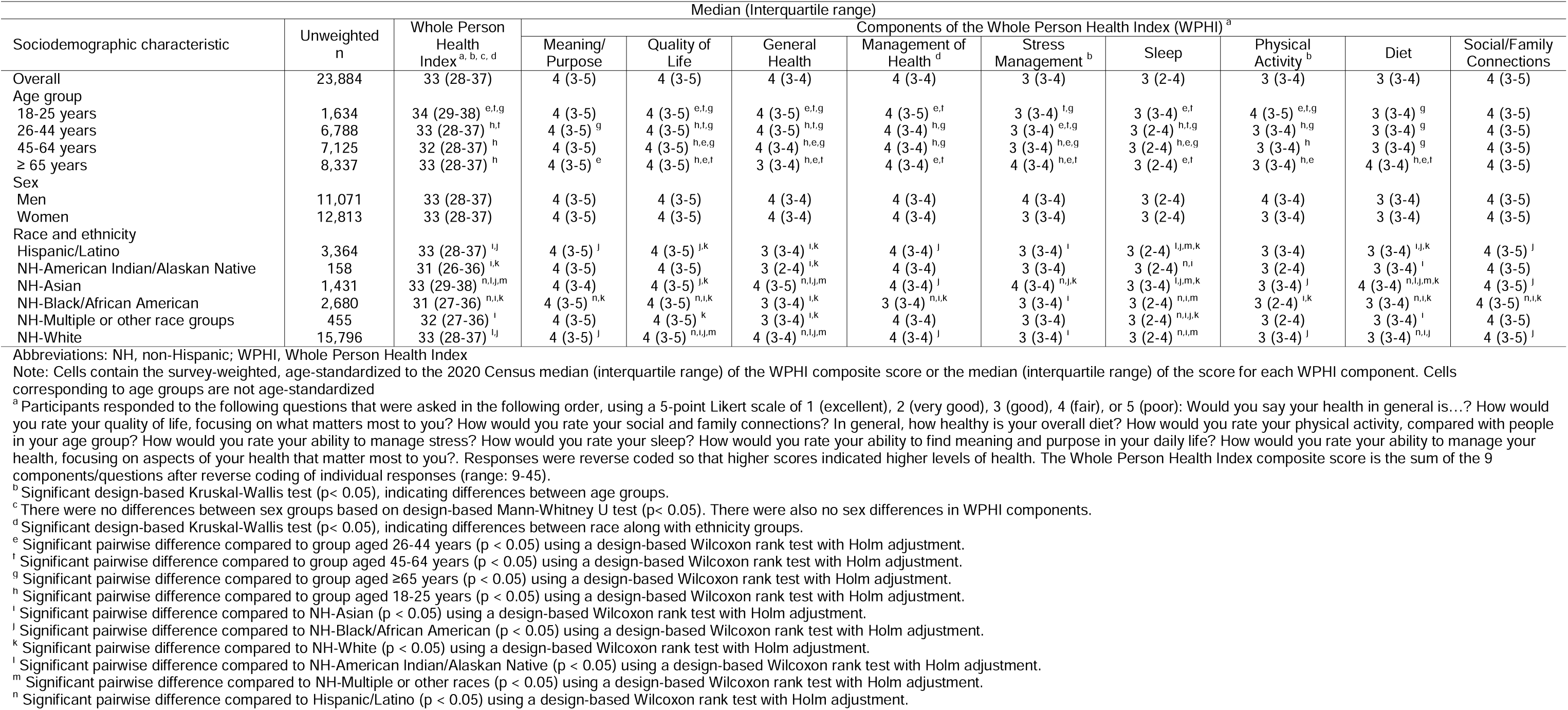
Medians and Interquartile Ranges of the Whole Person Health Index (WPHI) Composite Scores and WPHI Component Scores by Sociodemographic Characteristics, National Health Interview Survey, 2025 (N=23,884)

| Sociodemographic characteristic | Unweighted<br>n | Whole Person<br>Health<br>Index<sup>a, b, c, d</sup> | Median (Interquartile range) |  |  |  |  |  |  |  |  |
| --- | --- | --- | --- | --- | --- | --- | --- | --- | --- | --- | --- |
|  |  |  | Components of the Whole Person Health Index (WPHI)<sup>a</sup> |  |  |  |  |  |  |  |  |
|  |  |  | Meaning/<br>Purpose | Quality of<br>Life | General<br>Health | Management of<br>Health<sup>d</sup> | Stress<br>Management<sup>b</sup> | Sleep | Physical<br>Activity<sup>b</sup> | Diet | Social/Family<br>Connections |
| Overall | 23,884 | 33 (28-37) | 4 (3-5) | 4 (3-5) | 4 (3-4) | 4 (3-4) | 3 (3-4) | 3 (2-4) | 3 (3-4) | 3 (3-4) | 4 (3-5) |
| Age group |  |  |  |  |  |  |  |  |  |  |  |
| 18-25 years | 1,634 | 34 (29-38)<sup>e,f,g</sup> | 4 (3-5) | 4 (3-5)<sup>e,f,g</sup> | 4 (3-5)<sup>e,f,g</sup> | 4 (3-5)<sup>e,f</sup> | 3 (3-4)<sup>f,g</sup> | 3 (3-4)<sup>e,f</sup> | 4 (3-5)<sup>e,f,g</sup> | 3 (3-4)<sup>g</sup> | 4 (3-5) |
| 26-44 years | 6,788 | 33 (28-37)<sup>h,i</sup> | 4 (3-5)<sup>g</sup> | 4 (3-5)<sup>h,i,g</sup> | 4 (3-5)<sup>h,i,g</sup> | 4 (3-4)<sup>h,g</sup> | 3 (3-4)<sup>e,f,g</sup> | 3 (2-4)<sup>h,i,g</sup> | 3 (3-4)<sup>h,g</sup> | 3 (3-4)<sup>g</sup> | 4 (3-5) |
| 45-64 years | 7,125 | 32 (28-37)<sup>h</sup> | 4 (3-5) | 4 (3-5)<sup>h,e,g</sup> | 4 (3-4)<sup>h,e,g</sup> | 4 (3-4)<sup>h,g</sup> | 3 (3-4)<sup>h,e,g</sup> | 3 (2-4)<sup>h,e,g</sup> | 3 (3-4)<sup>h</sup> | 3 (3-4)<sup>g</sup> | 4 (3-5) |
| ≥ 65 years | 8,337 | 33 (28-37)<sup>h</sup> | 4 (3-5)<sup>e</sup> | 4 (3-5)<sup>h,e,f</sup> | 3 (3-4)<sup>h,e,f</sup> | 4 (3-4)<sup>e,f</sup> | 4 (3-4)<sup>h,e,f</sup> | 3 (2-4)<sup>e,f</sup> | 3 (3-4)<sup>h,e</sup> | 4 (3-4)<sup>h,e,f</sup> | 4 (3-5) |
| Sex |  |  |  |  |  |  |  |  |  |  |  |
| Men | 11,071 | 33 (28-37) | 4 (3-5) | 4 (3-5) | 4 (3-4) | 4 (3-4) | 4 (3-4) | 3 (2-4) | 4 (3-4) | 3 (3-4) | 4 (3-5) |
| Women | 12,813 | 33 (28-37) | 4 (3-5) | 4 (3-5) | 4 (3-4) | 4 (3-4) | 3 (3-4) | 3 (2-4) | 3 (3-4) | 3 (3-4) | 4 (3-5) |
| Race and ethnicity |  |  |  |  |  |  |  |  |  |  |  |
| Hispanic/Latino | 3,364 | 33 (28-37)<sup>i,j</sup> | 4 (3-5)<sup>j</sup> | 4 (3-5)<sup>i,k</sup> | 3 (3-4)<sup>i,k</sup> | 4 (3-4)<sup>j</sup> | 3 (3-4)<sup>i</sup> | 3 (2-4)<sup>i,j,m,k</sup> | 3 (3-4) | 3 (3-4)<sup>i,j,k</sup> | 4 (3-5)<sup>j</sup> |
| NH-American Indian/Alaskan Native | 158 | 31 (26-36)<sup>i,k</sup> | 4 (3-5) | 4 (3-5) | 3 (2-4)<sup>i,k</sup> | 4 (3-4) | 3 (3-4) | 3 (2-4)<sup>n,i</sup> | 3 (2-4) | 3 (3-4)<sup>i</sup> | 4 (3-5) |
| NH-Asian | 1,431 | 33 (29-38)<sup>n,i,j,m</sup> | 4 (3-4) | 4 (3-5)<sup>j,k</sup> | 4 (3-5)<sup>n,i,j,m</sup> | 4 (3-4)<sup>j</sup> | 4 (3-4)<sup>n,j,k</sup> | 3 (3-4)<sup>i,j,m,k</sup> | 3 (3-4)<sup>j</sup> | 4 (3-4)<sup>n,i,j,m,k</sup> | 4 (3-5)<sup>j</sup> |
| NH-Black/African American | 2,680 | 31 (27-36)<sup>n,i,k</sup> | 4 (3-5)<sup>n,k</sup> | 4 (3-5)<sup>n,i,k</sup> | 3 (3-4)<sup>i,k</sup> | 3 (3-4)<sup>n,i,k</sup> | 3 (3-4)<sup>i</sup> | 3 (2-4)<sup>n,i,m</sup> | 3 (2-4)<sup>i,k</sup> | 3 (3-4)<sup>n,i,k</sup> | 4 (3-5)<sup>n,i,k</sup> |
| NH-Multiple or other race groups | 455 | 32 (27-36)<sup>i</sup> | 4 (3-5) | 4 (3-5)<sup>k</sup> | 3 (3-4)<sup>i,k</sup> | 4 (3-4) | 3 (3-4) | 3 (2-4)<sup>n,i,j,k</sup> | 3 (2-4) | 3 (3-4)<sup>i</sup> | 4 (3-5) |
| NH-White | 15,796 | 33 (28-37)<sup>i,j</sup> | 4 (3-5)<sup>j</sup> | 4 (3-5)<sup>n,i,j,m</sup> | 4 (3-4)<sup>n,i,j,m</sup> | 4 (3-4)<sup>j</sup> | 3 (3-4)<sup>i</sup> | 3 (2-4)<sup>n,i,m</sup> | 3 (3-4)<sup>j</sup> | 3 (3-4)<sup>n,i,j</sup> | 4 (3-5)<sup>j</sup> |

Abbreviations: NH, non-Hispanic; WPHI, Whole Person Health Index
Note: Cells contain the survey-weighted, age-standardized to the 2020 Census median (interquartile range) of the WPHI composite score or the median (interquartile range) of the score for each WPHI component. Cells corresponding to age groups are not age-standardized
<sup>a</sup> Participants responded to the following questions that were asked in the following order, using a 5-point Likert scale of 1 (excellent), 2 (very good), 3 (good), 4 (fair), or 5 (poor): Would you say your health in general is...? How would you rate your quality of life, focusing on what matters most to you? How would you rate your social and family connections? In general, how healthy is your overall diet? How would you rate your physical activity, compared with people in your age group? How would you rate your ability to manage stress? How would you rate your sleep? How would you rate your ability to find meaning and purpose in your daily life? How would you rate your ability to manage your health, focusing on aspects of your health that matter most to you?. Responses were reverse coded so that higher scores indicated higher levels of health. The Whole Person Health Index composite score is the sum of the 9 components/questions after reverse coding of individual responses (range: 9-45).
<sup>b</sup> Significant design-based Kruskal-Wallis test ( $p < 0.05$ ), indicating differences between age groups.
<sup>c</sup> There were no differences between sex groups based on design-based Mann-Whitney U test ( $p < 0.05$ ). There were also no sex differences in WPHI components.
<sup>d</sup> Significant design-based Kruskal-Wallis test ( $p < 0.05$ ), indicating differences between race along with ethnicity groups.
<sup>e</sup> Significant pairwise difference compared to group aged 26-44 years ( $p < 0.05$ ) using a design-based Wilcoxon rank test with Holm adjustment.
<sup>f</sup> Significant pairwise difference compared to group aged 45-64 years ( $p < 0.05$ ) using a design-based Wilcoxon rank test with Holm adjustment.
<sup>g</sup> Significant pairwise difference compared to group aged ≥65 years ( $p < 0.05$ ) using a design-based Wilcoxon rank test with Holm adjustment.
<sup>h</sup> Significant pairwise difference compared to group aged 18-25 years ( $p < 0.05$ ) using a design-based Wilcoxon rank test with Holm adjustment.
<sup>i</sup> Significant pairwise difference compared to NH-Asian ( $p < 0.05$ ) using a design-based Wilcoxon rank test with Holm adjustment.

### Statistical analysis

We evaluated data missingness for the WPHI, its components, and core sociodemographic variables. Missing data patterns were evaluated using Stata’s *misstable* command suite. Logistic regression was used to estimate the association between a missingness indicator and core sociodemographic variables. We also used Little’s missing completely at random (MCAR) test to assess whether data were missing completely at random.

Survey weights that accounted for the sampling strategy, 11 months of sampling, and non-response; strata; and primary sampling units were then applied to all analyses to approximate nationally representative estimates. We described the study population with the weighted mean and standard error for continuous age and with unweighted counts, weighted proportions, and weighted, age-adjusted proportions (for all variables except age) for categorical variables. The 2020 US Census population served as the reference population for age-standardization, which accounted for differing age distributions across population subgroups (e.g., racial and ethnic groups). This approach was applied to account for age-related differences in WPHI;^19^ it also removed other differences that may emerge due to variation by age across groups.

Subsequent to describing the study sample, we assessed and determined a non-linear distribution of both the WPHI composite and individual WPHI item scores. Therefore, we estimated age-standardized (except for age), weighted medians (interquartile ranges [IQRs]) overall and by age, sex, and racial along with ethnic group. We then used omnibus Kruskal-Wallis tests for differences between more than two groups or omnibus Mann-Whitney U tests for differences between two groups. Post-hoc pairwise design-based Wilcoxon rank tests with Holm adjustment for multiple comparisons were then performed to compare each independent group.

We also determined the age-adjusted (except across age groups), weighted prevalence or proportion (95% confidence interval [CI]) of participants with an “optimal” WPHI composite score as well as “optimal” individual WPHI items/components. Rao-Scott χ² were used to test for differences in prevalence by age group, sex, and race along with ethnicity. We then estimated the pairwise survey-weighted contrasts of age-standardized (except for age groups) proportions of “optimal” WPH and WPH components with Holm adjustment for multiple comparisons. As a sensitivity analysis, we defined “optimal” WPHI composite and individual WPHI items scores using the threshold of ≥3 (good/very good/excellent) vs. ≤2 (fair/poor). Analyses were completed using survey procedures in Stata 19.5 (College Station, Texas), and a two-sided p-value <0.05 was used to determine statistical significance.

## RESULTS

### Missing data

The total missing across the WPHI, its components, and core demographic variables was low (∼1%). Results of logistic regression to evaluate relationships with missingness indicated that missingness was associated with age (OR = 1.02 [95% CI: 1.01–1.03]), suggesting these data were missing at random (MAR). Little’s MCAR test verified that these data were not missing completely at random (χ²(1169) = 1633.36, p < 0.001), thereby confirming MAR. We proceeded with a complete-case analysis, as listwise deletion under these conditions preserves statistical power, and adjusting for age can eliminate bias due to missing data ^20^. Survey design diagnostics (i.e., *svydescribe*) further confirmed that this approach did not compromise or result in empty primary sampling units or strata.

### Study population

Among 23,884 NHIS participants aged 18 years and older, representing approximately 261,620,640 US adults, mean age (SE) was 48 (0.2) years (Table 1). Approximately half (51%) were women, most identified as NH-White (60%), followed by Hispanic/Latino of any race (19%), NH-Black (12%), NH-Asian (6.6%), NH-multiple race groups or another race (2.0%), or NH-AIAN (0.6%).

### Whole person health, overall

#### Scores

Overall, the age-adjusted, survey-weighted median (IQR) WPHI score was 33 (28-37) (Table 2). The scores of individual WPHI components were highest for meaning/purpose, QOL, general health, management of health, and social/family connections (all medians = 4, corresponding to ‘very good’) and were lowest for stress management, sleep, physical activity, and diet (all medians = 3, corresponding to ‘good’).

#### “Optimal” (‘excellent’ or ‘very good’) WPH

Overall, 57% of adults (95% CI: 56%-58%) rated their WPH as ‘excellent’ or ‘very good,’ based on the average of their scores across the nine WPHI components (Table 3). Adults frequently rated their social/family connections (72% [95% CI: 72%-73%]) and QOL (70% [69%-70%]) as ‘excellent’ or ‘very good’. Fewer but more than half reported ‘excellent’ or ‘very good’ sense of meaning/purpose (62% [61%-62%], general health (56% [55%-57%]), and management of their personal health (55% [54%-56%]). Less than half reported ‘excellent’ or ‘very good’ stress management (49% [49%-50%]), physical activity compared to others their age (47% [46%-48%]), and diets (46% [45%-47%]). Comparatively fewer adults –slightly over one in three (36% [35%-37%])– reported their sleep as ‘excellent’ or ‘very good’.

**Table 3.** Prevalence of “optimal” whole person health and whole person health components by sociodemographic characteristics, National Health Interview Survey, 2025 (N=23,884)

| Sociodemographic characteristic | Unweighted n | Prevalence (95% CI) of an average response of 'excellent' or 'very good' to the Whole Person Health Index<sup>a, b, c</sup> | Prevalence (95% CI) of endorsement of 'excellent' or 'very good' to questions/components of the Whole Person Health Index (WPHI)<sup>a</sup> |  |  |  |  |  |  |  |  |
| --- | --- | --- | --- | --- | --- | --- | --- | --- | --- | --- | --- |
|  |  |  | Meaning/<br>Purpose<sup>b, c, d</sup> | Quality of Life<sup>b, c, d</sup> | General Health<sup>b, c</sup> | Management of Health<sup>b, c, d</sup> | Stress Management<sup>b, c, d</sup> | Sleep<sup>b, c, d</sup> | Physical Activity<sup>b, c, d</sup> | Diet<sup>b, c, d</sup> | Social/Family Connections<sup>c, d</sup> |
| Overall | 23,884 | 57.2<br>(56.3-58.1) | 61.6<br>(60.7-62.4) | 69.6<br>(68.8-70.4) | 55.7<br>(54.8-56.6) | 54.9<br>(54.0-55.8) | 49.3<br>(48.5-50.1) | 35.7<br>(34.9-36.5) | 47.1<br>(46.2-48.0) | 45.7<br>(44.8-46.6) | 72.2<br>(71.5-73.0) |
| Age group |  |  |  |  |  |  |  |  |  |  |  |
| 18-25 years | 1,634 | 57.2<br>(52.6-61.8) | 58.1<br>(53.5-62.7) | 69.6<br>(65.2-74.1) | 69.4<br>(65.2-73.7) | 56.0<br>(51.6-60.4) | 45.5<br>(40.9-50.2) | 38.6<br>(34.2-42.9) | 43.6<br>(39.6-47.7) | 45.1<br>(40.7-49.4) | 69.9<br>(65.7-74.0) |
| 26-44 years | 6,788 | 58.0<br>(56.6-59.4) | 63.4<br>(62.0-64.7) | 72.0<br>(70.7-73.3) | 62.8<br>(61.3-64.2) | 52.9<br>(51.5-54.3) | 47.4<br>(46.0-48.8) | 35.8<br>(34.4-37.3) | 44.7<br>(43.2-46.1) | 43.9<br>(42.4-45.3) | 72.3<br>(71.0-73.6) |
| 45-64 years | 7,125 | 55.2<br>(53.7-56.7) | 61.6<br>(60.1-63.1) | 68.0<br>(66.6-69.4) | 50.8<br>(49.2-52.3) | 53.3<br>(51.9-54.7) | 49.4<br>(48.0-50.7) | 33.5<br>(32.1-34.9) | 46.9<br>(45.4-48.3) | 44.5<br>(43.1-45.9) | 71.6<br>(70.3-73.0) |
| ≥ 65 years | 8,337 | 56.7<br>(55.4-58.0) | 59.5<br>(58.2-60.8) | 65.1<br>(63.7-66.4) | 42.9<br>(41.6-44.3) | 58.6<br>(57.2-59.9) | 53.9<br>(52.6-55.2) | 36.2<br>(34.9-37.5) | 48.9<br>(47.6-50.2) | 50.9<br>(49.5-52.2) | 72.8<br>(71.6-73.9) |
| Sex |  |  |  |  |  |  |  |  |  |  |  |
| Men | 11,071 | 57.7<br>(56.5-58.9) | 60.2<br>(59.0-61.4) | 68.1<br>(67.0-69.3) | 56.0<br>(54.8-57.1) | 53.9<br>(52.7-55.1) | 53.6<br>(52.5-54.8) | 37.4<br>(36.3-38.6) | 52.2<br>(51.1-53.4) | 44.7<br>(43.5-45.8) | 70.5<br>(69.4-71.5) |
| Women | 12,813 | 56.7<br>(55.5-57.9) | 62.9<br>(61.8-64.0) | 71.0<br>(69.9-72.0) | 55.4<br>(54.1-56.6) | 55.8<br>(54.6-57.0) | 45.2<br>(44.1-46.2) | 34.1<br>(33.1-35.1) | 42.1<br>(41.0-43.3) | 46.8<br>(45.6-48.0) | 73.9<br>(72.8-74.9) |
| Race and ethnicity |  |  |  |  |  |  |  |  |  |  |  |
| Hispanic/Latino | 3,364 | 55.7<br>(53.4-58.0) | 60.2<br>(57.9-62.5) | 65.1<br>(63.0-67.2) | 48.0<br>(45.9-50.1) | 53.0<br>(50.9-55.0) | 48.6<br>(46.5-50.8) | 39.0<br>(36.8-41.1) | 46.0<br>(43.8-48.1) | 43.2<br>(41.0-45.3) | 71.4<br>(69.3-73.4) |
| NH-American Indian/Native Alaskan | 158 | 47.7<br>(38.5-56.9) | 55.1<br>(44.7-65.5) | 64.9<br>(55.0-74.9) | 46.9<br>(36.9-57.0) | 52.6<br>(42.8-62.4) | 48.0<br>(37.1-58.8) | 27.0<br>(17.5-36.5) | 44.4<br>(35.7-53.1) | 37.1<br>(27.5-46.7) | 62.0<br>(54.4-69.7) |
| NH-Asian | 1,431 | 61.4<br>(58.3-64.5) | 60.4<br>(57.2-63.6) | 66.8<br>(63.7-69.9) | 59.8<br>(56.5-63.0) | 56.5<br>(53.0-59.9) | 53.5<br>(50.3-56.6) | 40.6<br>(37.5-43.6) | 48.7<br>(45.3-52.1) | 57.2<br>(54.0-60.4) | 74.9<br>(72.3-77.5) |
| NH-Black/African American | 2,680 | 47.6<br>(45.2-49.9) | 54.3<br>(51.8-56.7) | 60.1<br>(57.7-62.5) | 46.3<br>(44.0-48.5) | 48.4<br>(46.1-50.8) | 46.0<br>(43.6-48.4) | 34.2<br>(31.9-36.5) | 42.2<br>(39.9-44.6) | 35.9<br>(33.7-38.2) | 65.0<br>(62.9-67.1) |
| NH-Multiple or other race groups | 455 | 51.2<br>(46.1-56.3) | 62.3<br>(57.1-67.5) | 65.5<br>(60.8-70.3) | 48.5<br>(43.7-53.3) | 51.4<br>(46.4-56.5) | 48.2<br>(43.2-53.3) | 28.8<br>(24.0-33.6) | 44.1<br>(38.8-49.5) | 41.2<br>(35.8-46.5) | 67.1<br>(62.2-72.1) |
| NH-White | 15,796 | 59.0<br>(57.9-60.1) | 63.5<br>(62.5-64.5) | 73.0<br>(72.1-74.0) | 59.4<br>(58.2-60.5) | 56.4<br>(55.3-57.4) | 49.6<br>(48.6-50.6) | 34.6<br>(33.6-35.6) | 48.3<br>(47.1-49.4) | 47.4<br>(46.3-48.5) | 73.8<br>(72.8-74.7) |

Abbreviations: CI, Confidence Interval; NH, non-Hispanic; WPHI, Whole Person Health Index
Note: Cells contain the survey-weighted, age-standardized to the 2020 Census prevalence or proportion (95% confidence interval) of individuals responding 'excellent' or 'very good' (vs. 'good'/'fair'/'poor').
<sup>a</sup> Participants responded to the following questions that were asked in the following order, using a 5-point Likert scale of 1 (excellent), 2 (very good), 3 (good), 4 (fair), or 5 (poor): Would you say your health in general is...? How would you rate your quality of life, focusing on what matters most to you? How would you rate your social and family connections? In general, how healthy is your overall diet? How would you rate your physical activity, compared with people in your age group? How would you rate your ability to manage stress? How would you rate your sleep? How would you rate your ability to find meaning and purpose in your daily life? How would you rate your ability to manage your health, focusing on aspects of your health that matter most to you? Responses were reverse coded so that higher scores indicated higher levels of health; scores were then dichotomized as ≥4 ('excellent' or 'very good') vs. '≤3 'good', 'fair', or 'poor'. To estimate average whole person health, the nine individual items were averaged, rounded to the nearest integer and dichotomized as ≥4 ('excellent' or 'very good') vs. '≤3 'good', 'fair', or 'poor'.
<sup>b</sup> p-value for the omnibus Rao-Scott p-value <0.05, indicating differences between age groups.
<sup>c</sup> p-value for the omnibus Rao-Scott p-value <0.05, indicating differences between race and ethnicity groups.
<sup>d</sup> p-value for the omnibus Rao-Scott p-value <0.05, indicating differences between sex groups.

### Whole person health by age, sex, and race along with ethnicity

Both WPHI scores and prevalence of “optimal” WPH (rated as ‘excellent’ or ‘very good’), based on the average of their scores across the nine WPHI components, varied by age and race along with ethnicity (Tables 2-4).

**Table 4.** Pairwise comparisons of the prevalence of “optimal” whole person health by sociodemographic characteristics, National Health Interview Survey, 2025 (N=23,884)

| Sociodemographic characteristic | Pairwise comparison | Prevalence difference (95% CI) of an average response of 'excellent' or 'very good' to the Whole Person Health Index <sup>a</sup> | p-value | Holm adjusted p-value |
| --- | --- | --- | --- | --- |
| Age group | 18-25 years vs. 26-44 years | 2.9 (-0.1 to 6.0) | 0.060 | 0.180 |
|  | 18-25 years vs. 45-64 years | 5.7 (2.7 to 8.8) | 0.000 | 0.002 |
|  | 18-25 years vs. ≥65 years | 4.2 (1.3 to 7.2) | 0.005 | 0.025 |
|  | 26-44 years vs. 45-64 years | 2.8 (0.8 to 4.8) | 0.006 | 0.025 |
|  | 26-44 years vs. ≥65 years | 1.3 (-0.5 to 3.1) | 0.163 | 0.218 |
|  | 45-64 years vs. ≥65 years | -1.5 (-3.3 to 0.3) | 0.109 | 0.218 |
| Race and ethnicity | Hispanic/Latino vs. NH-American Indian/Alaskan Native | 9.2 (-0.4 to 18.9) | 0.061 | 0.368 |
|  | Hispanic/Latino vs. NH-Asian | -5.1 (-8.8 to -1.5) | 0.006 | 0.061 |
|  | Hispanic/Latino vs. NH-Black/African American | 8.5 (5.2 to 11.9) | 0.000 | 0.000 |
|  | Hispanic/Latino vs. NH-Multiple or other race groups | 3.3 (-2.2 to 8.9) | 0.241 | 0.724 |
|  | Hispanic/Latino vs. NH-White | -2.5 (-4.9 to -0.1) | 0.040 | 0.282 |
|  | NH-American Indian/Alaskan Native vs. NH-Asian | -14.4 (-24.1 to -4.7) | 0.004 | 0.046 |
|  | NH-American Indian/Alaskan Native vs. NH-Black/African American | -0.7 (-10.4 to 9.0) | 0.890 | 0.890 |
|  | NH-American Indian/Alaskan Native vs. NH-Multiple or other race groups | -5.9 (-16.7 to 4.9) | 0.285 | 0.724 |
|  | NH-American Indian/Alaskan Native vs. NH-White | -11.7 (-21.3 to -2.1) | 0.017 | 0.149 |
|  | NH-Asian vs. NH-Black/African American | 13.7 (9.8 to 17.6) | 0.000 | 0.000 |
|  | NH-Asian vs. NH-Multiple or other race groups | 8.5 (2.5 to 14.4) | 0.006 | 0.061 |
|  | NH-Asian vs. NH-White | 2.6 (-0.6 to 5.8) | 0.109 | 0.435 |
|  | NH-Black/African American vs. NH-Multiple or other race groups | -5.2 (-10.8 to 0.4) | 0.069 | 0.368 |
|  | NH-Black/African American vs. NH-White | -11.0 (-13.7 to -8.4) | 0.000 | 0.000 |
|  | NH-Multiple or other race groups vs. NH-White | -5.8 (-11.0 to -0.7) | 0.027 | 0.215 |

Abbreviations: CI, Confidence Interval; NH, non-Hispanic; WPHI, Whole Person Health Index
Note: Results show the survey-weighted contrasts of age-standardized (except for age groups) proportions with Holm adjustment for multiple comparisons. Comparisons are of individuals responding 'excellent' or 'very good' (vs. 'good'/'fair'/'poor') within each group.
<sup>a</sup>Participants responded to the following questions that were asked in the following order, using a 5-point Likert scale of 1 (excellent), 2 (very good), 3 (good), 4 (fair), or 5 (poor): Would you say your health in general is...? How would you rate your quality of life, focusing on what matters most to you? How would you rate your social and family connections? In general, how healthy is your overall diet? How would you rate your physical activity, compared with people in your age group? How would you rate your ability to manage stress? How would you rate your sleep? How would you rate your ability to find meaning and purpose in your daily life? How would you rate your ability to manage your health, focusing on aspects of your health that matter most to you?. Responses were reverse coded so that higher scores indicated higher levels of health. To estimate average whole person health, the nine individual items were averaged, rounded to the nearest integer, and dichotomized as ≥4 ('excellent' or 'very good') vs. ≤3 ('good', 'fair', or 'poor').

#### Age

WPHI score was highest among adults aged 18-25 years (median (IQR)=43 [29-38]; Table 2). Prevalence of “optimal” WPH was lowest among adults aged 45-64 years (55% [54%-57%]) and highest among adults aged 26-44 years (58% [57%-59%]) (Table 3). Post-hoc pairwise comparisons (Table 4) revealed higher prevalence of “optimal” WPH among both 18– 25-year-olds (prevalence difference (PD)=5.7 [95% CI: 2.7,8.8]) and 26–44-year-olds (PD=2.8 [0.8,4.8]) compared with 45–64-year-olds. Adults aged 18-25 years also had higher prevalence of “optimal” WPH compared with adults aged ≥65 years (PD=4.2 [1.3,7.2]). There were no other differences in WPH by age.

#### Sex

Neither WPHI scores nor prevalence of optimal WPH varied by sex (e.g., prevalence of “optimal” WPH men = 58% [95% CI:57%-59%], women = 57% [56%-58%]; Tables 3).

#### Race and ethnicity

WPHI scores varied across racial and ethnic groups, and prevalence of “optimal” WPH was lowest among NH-American Indian/Native Alaskan (48% [39%-57%]) and NH-Black/African American adults (48% [45%-50%]) (Tables 2-3). In post-hoc comparisons with Holm adjustment for multiple comparisons (Table 4), NH-Black/African American adults had a lower prevalence of “optimal” WPH compared to Hispanic/Latino, NH-Asian, and NH-White adults. Compared to NH-Black adults, prevalence of “optimal” WPH was 8.5 (95% CI: 5.2,12) and 14 (95% CI: 9.8,18) percentage points higher among Hispanic/Latino and NH-Asian adults, respectively; NH-Black adults had 11 percentage points (95% CI: -14,-8.4) lower prevalence of “optimal” WPH compared to NH-White adults.

### Components of whole person health by age, sex, and race along with ethnicity

#### Age

Scores of WPHI components and the prevalence of “optimal” (i.e., rating ‘excellent’ or ‘very good’) WPH components differed by age between at least two age groups for all except social and family connections (p<0.05 for design-based Wilcoxon rank sum tests with Holm adjustment; Tables 2-3); however, patterns in pairwise differences varied by component. Only adults aged 26-44 years and 45-64 years differed in meaning and purpose; optimal prevalence was higher among 26-44 year-olds. Younger age groups had a higher prevalence of reporting ‘excellent’ or ‘very good’ QOL and general health than each older age group (Figure 1). Differences were largest for general health (e.g., 18-25 year olds vs. ≥65 year olds: QOL PD=9.9 [95% CI:7.4, 12.5]; general health PD=29 [27, 32]). The prevalence of “optimal” management of health was lower among adults aged 26-44 years and 45-64 years compared with both the youngest (18-25 years) and oldest (≥65 years) age groups. The oldest age group also had the highest prevalence of “optimal” stress management and diet compared to all the younger age groups. While the prevalence of “optimal” sleep was higher among 18-25 year olds compared to all older age groups, adults aged 45-64 years were less likely to report “optimal” sleep than adults aged ≥65 years. Compared to adults aged ≥65 years, the prevalence of “optimal” physical activity was higher among 18–25-year-olds but lower among adults aged 26-44 years.

**Figure 1.**
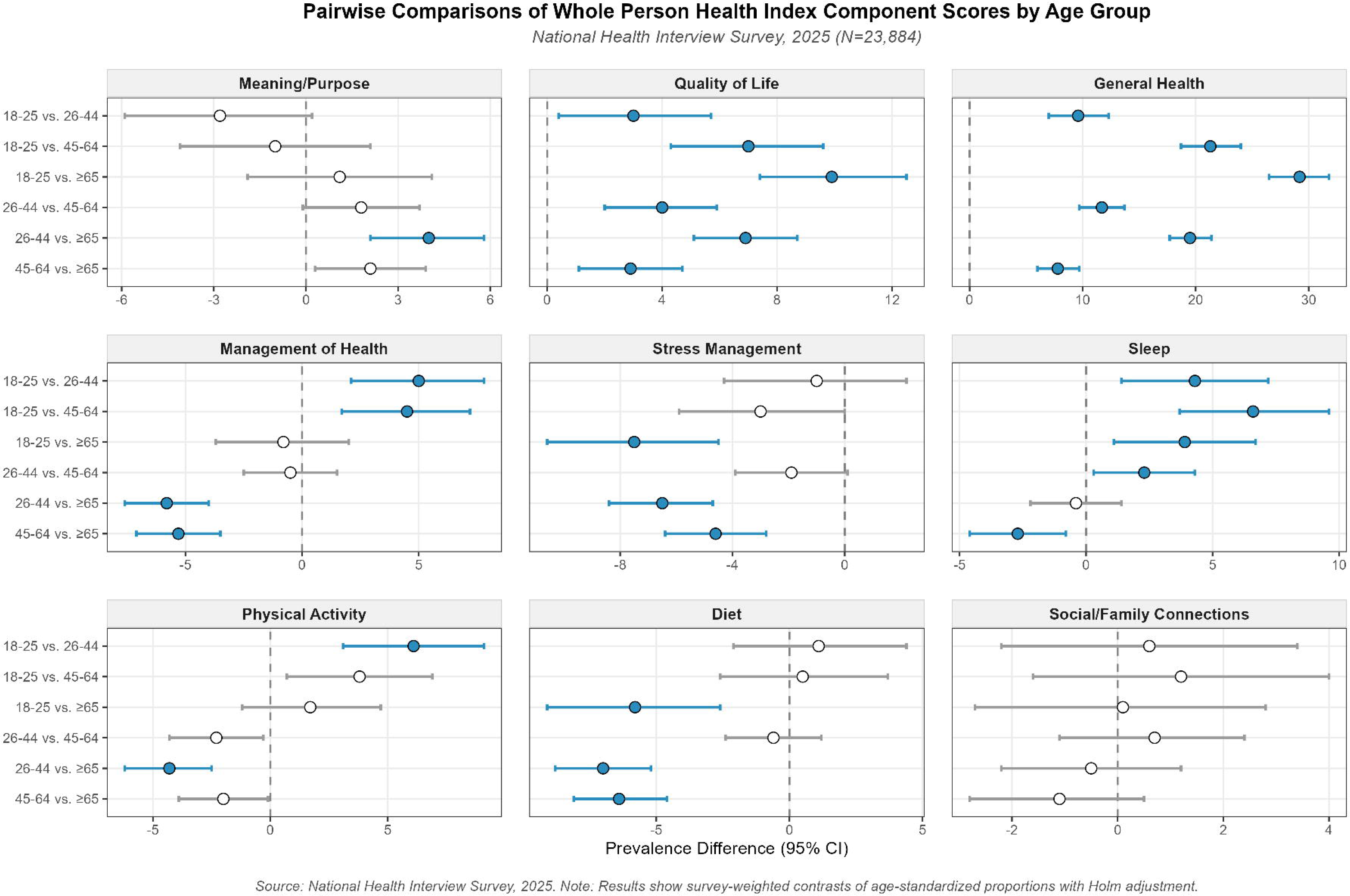
Pairwise comparisons of the prevalence of “optimal” whole person health components by age. Prevalence differences of survey-weighted proportions and 95% confidence intervals are shown. Color indicates statistical significance after Holm adjustment. Source: National Health Interview Survey, 2025 (N=23,884).

#### Sex

Although scores for the nine WPHI components did not vary by sex, women were more likely than men to report ‘excellent’ or ‘very good’ sense of meaning or purpose, QOL, management of health, diet, and social and family connections (Tables 2-3). Conversely, men were more likely than women to report ‘excellent’ or ‘very good’ management of stress, sleep, and physical activity. Men and women did not differ in rating their perceived general health.

#### Race and ethnicity

Across all WPHI components, the scores and the prevalence of reporting ‘excellent’ or ‘very good’ (i.e., “optimal” values) varied between at least two racial and ethnic groups after age-adjustment and Holm adjustment for multiple comparisons (Tables 2-3, Figure 2). Prevalence of “optimal” health across WPHI components was generally highest among NH-Asian and NH-White adults and lowest among either NH-AIAN or NH-Black adults, and the largest differences were in general health (e.g., PD_NH-Asian vs. NH-Black_=13 [95% CI: 9.1, 17]), sleep (e.g., PD_NH-AIAN vs. NH-Asian_=-15 [-25, -4.7]), diet (e.g., PD_NH-AIAN vs. NH-Asian_=-22 [-32, -12]), and social/family connections. Hispanic/Latino adults also often had lower prevalence of “optimal” WPHI components compared with NH-Asian and NH-White adults but higher prevalence than NH-Black adults (e.g., QOL, diet). For instance, while 62% of NH-AIAN adults and 65% of NH-Black adults perceived ‘excellent’ or ‘very good’ social/family connections, ‘excellent’ or ‘very good’ social/family connections was reported by 71% of Hispanic/Latino adults, 74% of NH-White adults, and 75% of NH-Asian adults (see Supplemental Table 2 for CIs).

**Figure 2.**
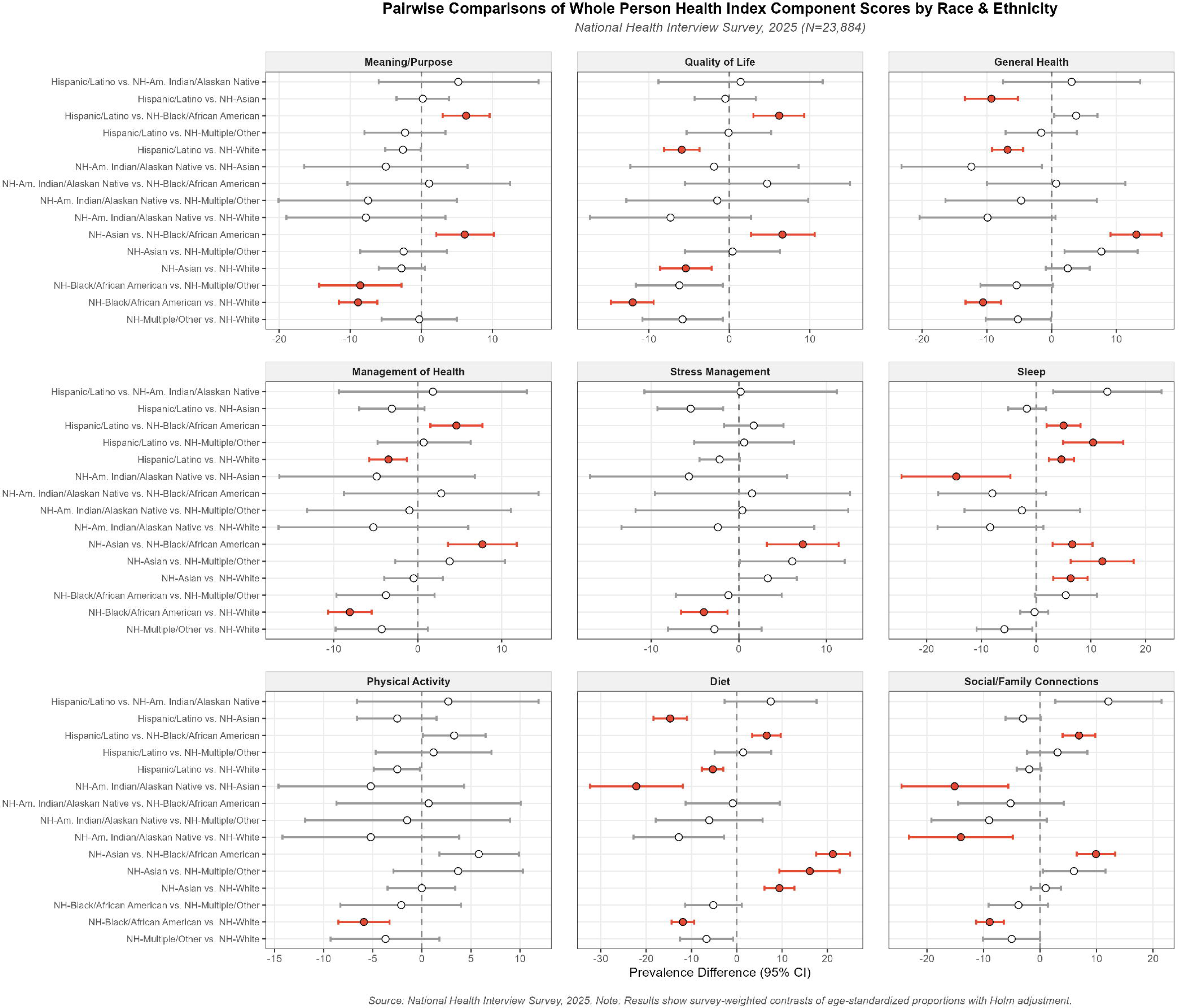
Pairwise comparisons of the prevalence of “optimal” whole person health components by race and ethnicity. Prevalence differences of survey-weighted, age-adjusted proportions and 95% confidence intervals are shown. Color indicate statistical significance after Holm adjustment. Source: National Health Interview Survey, 2025 (N=23,884).

### Sensitivity analysis

When considering ‘excellent’, ‘very good’, or ‘good’ as the “optimal” threshold for WPH, 93% of adults (95% CI:92%-93%) met criteria for “optimal” WPH based on the average of their scores across the nine WPHI components (Supplemental Table 3). Only pairwise differences in “optimal” WPH by race and ethnicity remained, the pattern was consistent with the main analysis: prevalence among NH-Black adults (88% [84%-91%]) was lower compared to NH-Asian (95% [94%-97%]) and NH-White (93% [93%-94%]) adults after adjustment for multiple comparisons (Supplemental Tables 3-4). Further, prevalence of “optimal” WPH was now higher among NH-Asian adults compared to all other racial and ethnic groups (range: 2.3 [vs. NH-White] to 6.5 [vs. NH-multiple or other race groups] percentage point difference). For WPHI components (Supplemental Table 5), there were no longer age differences in meaning and purpose or physical activity. However, patterns by age for the remaining WPHI components were consistent with the main analysis. Although attenuated, “optimal” QOL, diet, and social/family connections remained more prevalent among women, and “optimal” stress management, sleep, and physical activity remained more prevalent among men. By race and ethnicity, prevalence of “optimal” WPHI components remained lowest among NH-AIAN and NH-Black adults, especially in comparison to NH-Asian adults, who generally had the highest prevalence of “optimal” values across most components followed by NH-White adults.

## DISCUSSION

In this descriptive study of whole person health (WPH) and its components– encompassing spiritual, mental, physical, behavioral, and social health domains–slightly over half of US adults (57%) reported, on average, ‘excellent’ or ‘very good’ WPH (median WPHI score=33 [IQR: 28-37]). Across the WPH domains, US adults most often reported strong social relationships and subjective wellbeing, as reflected in the high prevalence of ‘excellent’ or ‘very good’ quality of life (QOL) and social/family connections. In contrast, health behaviors received the lowest endorsements, particularly sleep. These findings indicate that health behaviors represent an important area for improvement towards WPH.

Nuances in WPH emerged by age, sex, and race along with ethnicity. Patterns by age differed across WPH and its individual components. Young adults had the highest prevalence of ‘excellent’ or ‘very good’ (which we considered “optimal”) overall WPH; however, age-related patterns were not uniform across domains. For example, older adults (≥65 years) more frequently reported ‘excellent’ or ‘very good’ stress management and diet. While overall WPH did not differ by sex, sex-differences were evident across WPH domains. Women more frequently endorsed ‘excellent’ or ‘very good’ ability to find meaning/purpose in life, QOL, health management, diet, and social connections while men more frequently endorsed ‘excellent’ or ‘very good’ stress management, sleep, and physical activity. These findings suggest that similar overall WPH may reflect different patterns of strengths across WPH domains. Consistent differences emerged across racial and ethnic groups. NH-AIAN and NH-Black adults had the lowest prevalence of “optimal” WPH overall and across WPH domains, NH-Asian and NH-White adults had the highest prevalence, and Hispanic/Latino adults generally fell between these groups. These patterns persisted and, for non-Hispanic Asian adults, became more pronounced in sensitivity analyses using a lower threshold to define “optimal” WPH, suggesting that the observed differences were robust to WPH threshold choice. Overall, these results indicate that both WPH and specific WPH domains warrant improvement. They also identify behavioral health, particularly sleep, as an important target for interventions while providing population-level targets for improving WPH.

The nuance and non-linearity in patterning of WPH components by age is consistent with established theoretical and empirical literature. “Optimal” overall WPH as well as QOL, general health, sleep, and physical activity among the youngest age group is likely explained by increases with age in chronic conditions,^2^ including pain,^21^ sleep disturbances,^22^ and functional limitations,^23^ all of which can impact multiple aspects of health and wellbeing.^24^ In contrast, reports of better stress management among adults in the oldest age groups are likely explained by the socioemotional selectivity theory as well as differences in stressor exposure, stress appraisal, and motivation.^25,26^ As people age, individuals are hypothesized to increasingly prioritize present-oriented, emotionally meaningful goals, a shift linked to emotional regulation and wellbeing later in life.^25,26^ However, emotional regulation also appears to remain relatively stable across adulthood.^27^ Older adults may experience fewer and less severe daily stressors, such as job stress, increasing perception of stress management. Better diet among older adults is consistent with documented age differences in dietary patterns observed with national surveillance data.^28,29^ A possible explanation is that fewer competing external demands such as work, caregiving, or social obligations allow older versus adults to better establish routines and meet nutritional needs, which has been linked to improved aging.^30^

Observed sex differences in WPH components also align with prior literature and may be explained by biopsychosocial pathways. The WPHI component-level findings are consistent with prior literature: women report higher diet quality, more sleep disturbances, and stronger social ties, while men report more leisure-time physical activity.^31–36^ Roles, such as caregiving and household responsibilities, can yield stronger relational engagement^37^ and are associated with more control over health management (e.g., health appointments, diet)^38^ but can result in less time for to engage in health behaviors, such as sleep, and poorer stress management among women.^32,34,39^ In contrast, more structured, work-based routines among men may support engagement in physical activity and thereby support stress management. Further, sex-specific patterns may be reinforced by hormonal factors that contribute to heightened stress reactivity and dysregulation of stress responses among women.^40,41^

Consistent racial and ethnic differences in both WPH and its components also align with prior component-specific evidence and are likely best explained by environmental and social factors. For instance, studies have shown that NH-AIAN and NH-Black adults generally have poorer sleep health,^34,42^ more food insecurity and lower diet quality,^43^ and more exposure to environmental hazards and social conditions that can negatively impact multiple WPH domains.^44,45^ Although Hispanic/Latino adults also face comparable socioeconomic constraints as NH-AIAN and NH-Black adults,^46^ Hispanic/Latino adults generally demonstrate more favorable social support and health outcomes.^47–49^ NH-Asian and NH-White adults generally have the highest socioeconomic status, and there is a well-established socioeconomic gradient of health.^50,51^ Therefore, differences across these groups are likely explained by differential socioeconomic, environmental, and social factors that can become biologically embedded and impact health behaviors as well as mental, physical, and social health through chronic stress pathways.^12,50,52^

Results should be interpreted in the context of limitations. Data were cross-sectional; however, the investigated sociodemographic characteristics, such as age, sex, and race along with ethnicity precede WPH, limiting concerns about temporality.^53^ All WPH data were self-reported, introducing potential bias related to differences in interpretations of the questions, which may have led to differential reporting. For instance, and consistent with the current study, prior evidence suggests that younger versus older adults with equivalent levels of WPH assess their general health more positively. We addressed this – at least in part –by applying age-adjustment throughout sex-and race and ethnicity-stratified analyses. But differences across age groups, particularly for general health, deserve the most cautious interpretations. Further, individual WPHI items or components should be interpreted cautiously as the WPHI index was designed to assess a single construct of WPH, and there are other standardized, validated instruments that more thoroughly capture each sub-construct within the WPHI.^19^ Additional studies comparing WPHI item scores to validated instruments, such as the Pittsburgh Sleep Quality Index^54^ for sleep can provide further insight about concordance between WPHI item scores and such instruments.^19^ Further, there is no standard threshold used to determine “optimal” values of the WPHI. While we averaged scores across individuals’ items and determined which individuals had “optimal” WPH, on average, this was likely a simplification of WPH as individual items are likely not equally impactful on health. For instance, a sense of meaning and purpose may precede health behaviors and have a stronger effect on achieving WPH than, for instance, physical activity. While our results can generate future hypotheses, validation and psychometric studies are necessary to determine what constitutes “optimal” WPH. Lastly, there is no standard definition of WPH,^9^ and other aspects of health that people may deem important for their overall health and wellbeing, such as sexual health, were not captured in the WPHI.^55^ This study was strengthened by the large nationally representative sample and its design. We assessed missing data and determined data to be MAR, reducing the likelihood that complete case analysis resulted in biased estimates. We adjusted for multiple comparisons, reducing the likelihood that results were due to chance.

## CONCLUSION

In this study of US adults in 2025, slightly more than half of the participants perceived their whole person health (WPH) across spiritual, mental/emotional, physical, behavioral, and social domains as ‘excellent’ or ‘very good’. The largest strengths were in social connections and the largest weaknesses were in health behaviors, particularly sleep. Patterns in WPH varied by age, sex, and racial and ethnic group. Older adults reported better diets and stress management, while younger adults reported better health across the remaining domains. Although men and women reported similar overall WPH, their domain-specific strengths differed, indicating that overall WPH may mask sex differences in achieving “optimal” health across individual domains. In particular, women were more likely than men to report ‘excellent’ or ‘very good’ sense of meaning or purpose, QOL, management of health, diet, and social and family connections. Men were more likely than women to report ‘excellent’ or ‘very good’ management of stress, sleep, and physical activity while their perceived general health did not differ. Furthermore, NH-AIAN and NH-Black adults were generally least likely to report “optimal” WPH and WPH components, whereas NH-Asian and NH-White adults were generally most likely to do so. These observed differences provide opportunities for hypothesis generation in future studies and identify potential domain – and population-specific intervention targets. Comprehensive systems approaches that create environments in which individuals can flourish may help improve health for all.

## Data Availability

All data produced in the present study are available upon reasonable request to the authors.

## Notes

Conflicts of interest: None declared

Sources of funding: This work was supported in part by the Intramural Research Program of the NIH, National Institute of Environmental Health Sciences (Z1AES103325 (CLJ)).

### Competing Interest Statement

The authors have declared no competing interest.

### Author Declarations

This analysis of secondary, deidentified data is not considered human subjects research; therefore, approval by the National Institutes of Health Institutional Review Board was waived. https://www.cdc.gov/nchs/nhis/documentation/2025-nhis.html#cdc_data_surveillance_section_2-questionnaires-and-survey-materials

