## Supplemental Material for "Whole person health across sociodemographic groups of adults in the United States"

Symielle A. Gaston, PhD, MPH ^1^

Christopher Payne, MA ^2^

Chandra L. Jackson, PhD, MS ^1,3^

^1^ Epidemiology Branch, National Institute of Environmental Health Sciences, National Institutes of Health, Department of Health and Human Services, Research Triangle Park, NC, USA

^2^ DLH, LLC, Bethesda, MD, USA

^3^	Intramural Research Program, National Institute on Minority Health and Health Disparities, National Institutes of Health, Department of Health and Human Services, Bethesda, MD, USA

Please direct correspondence to Chandra L. Jackson, PhD, MS at 111 TW Alexander Drive, Research Triangle Park, N.C. 27709; telephone: 984-287-3701; fax: 301-480-3290;.

Running Title:	Whole Person Health among US Adults

Conflicts of interest:	None declared

Sources of funding: This work was supported in part by the Intramural Research Program of the NIH, National Institute of Environmental Health Sciences (Z1AES103325 (CLJ)). The contributions of the NIH authors are considered Works of the United States Government. The findings and conclusions presented in this paper are those of the authors and do not necessarily reflect the views of the NIH or the U.S. Department of Health and Human Services.

The datasets generated during and/or analyzed during the current study are publicly available.

**CRediT:**

**Authors:** Symielle A. Gaston, Christopher Payne, Chandra L. Jackson

SAG contributed to methodology, project administration, writing- original draft, and writing – review & editing. CP contributed to methodology, formal analysis, visualization, validation, writing – original draft, and writing – review & editing. CLJ conceptualized the study, provided resources and supervision, and contributed to methodology and writing – review & editing.

### Supplemental Figure 1. Flow chart of study population selection

Final analytic sample

**(N=23,884)**

Total NHIS Sample,

2025

**(N=30,264)**

- Missing data
  - Whole Person Health Index (n=250)
  - Age (n=57)
  - Sex (n=24)
  - Race and Ethnicity (n=0)
- **Total excluded (n=331)**
- Age <18 years (n=6,049)

NHIS Sample of adults,

2025

**(N=24,215)**

### Supplemental Table 1. Comparison of included and excluded study participants, National Health Interview Survey, 2025 (N=24,215)

|  | Included n=23884 (98.6%) | Excluded n=331 (1.4%) | Chi-square or  t-test p-value |
| --- | --- | --- | --- |
| Characteristics ^a^ |  |  |  |
| Age (years), mean (SE) | 48.2 (0.2) | 55.2 (1.4) | <0.001 |
| Age group |  |  | <0.001 |
| 18-25 years | 13.4 | 6.6 |  |
| 26-44 years | 32.7 | 22.5 |  |
| 45-64 years | 30.8 | 25.0 |  |
| ≥65 years | 23.1 | 30.0 |  |
| Don't know/refused | 0.0 | 15.9 |  |
| Sex |  |  | <0.001 |
| Men | 48.8 | 46.1 |  |
| Women | 51.2 | 43.9 |  |
| Don't know/refused | 0.0 | 10.0 |  |
| Race and ethnicity |  |  | 0.310 |
| Hispanic/Latino | 18.7 | 18.9 |  |
| NH-American Indian/Alaskan Native | 0.6 | 0.6 |  |
| NH-Asian | 6.6 | 8.8 |  |
| NH-Black/African American | 12.0 | 16.1 |  |
| NH-Multiple or other race groups | 2.1 | 0.4 |  |
| NH-White | 60.1 | 55.2 |  |
| Don't know/refused | 0.0 | 0.0 |  |
| Abbreviations: SE, standard error; NH, non-Hispanic | | | |
| Note: All estimates are weighted for the survey's complex sampling design. Data are presented as column percentages or means and standard errors. Percentages may not sum to 100 due to rounding | | | |
| ^a^ Estimates except for age are age-standardized to the US 2020 population | | | |

### Supplemental Table 2. Pairwise comparisons of the prevalence of “optimal” whole person health components by sociodemographic characteristics,, National Health Interview Survey, 2025 (N=23,884)

| Sociodemographic characteristic | Pairwise comparison | Endorsement of 'excellent' or 'very good' to questions/components of the Whole Person Health Index (WPHI) ^a^ | | | | | | | | | | | | | | | | | | | | | | | | | | |
| --- | --- | --- | --- | --- | --- | --- | --- | --- | --- | --- | --- | --- | --- | --- | --- | --- | --- | --- | --- | --- | --- | --- | --- | --- | --- | --- | --- | --- |
|  |  | Meaning/Purpose | | | Quality of Life | | | General Health | | | Management of Health | | | Stress Management | | | Sleep | | | Physical Activity | | | Diet | | | Social/Family Connections | | |
|  |  | Prevalence difference (95% CI) | p-value | Holm adjusted p-value | Prevalence difference (95% CI) | p-value | Holm adjusted p-value | Prevalence difference (95% CI) | p-value | Holm adjusted p-value | Prevalence difference (95% CI) | p-value | Holm adjusted p-value | Prevalence difference (95% CI) | p-value | Holm adjusted p-value | Prevalence difference (95% CI) | p-value | Holm adjusted p-value | Prevalence difference (95% CI) | p-value | Holm adjusted p-value | Prevalence difference (95% CI) | p-value | Holm adjusted p-value | Prevalence difference (95% CI) | p-value | Holm adjusted p-value |
| Age group |  |  |  |  |  |  |  |  |  |  |  |  |  |  |  |  |  |  |  |  |  |  |  |  |  |  |  |  |
|  | 18-25 years vs. 26-44 years | -2.8 (-5.9 to 0.2) | 0.067 | 0.254 | 3.0 (0.4 to 5.7) | 0.026 | 0.026 | 9.6 (7.0 to 12.3) | 0.000 | 0.000 | 5.0 (2.1 to 7.8) | 0.001 | 0.003 | -1.0 (-4.3 to 2.2) | 0.532 | 0.532 | 4.3 (1.4 to 7.2) | 0.004 | 0.018 | 6.1 (3.1 to 9.1) | 0.000 | 0.000 | 1.1 (-2.1 to 4.4) | 0.489 | 1.000 | 0.6 (-2.2 to 3.4) | 0.697 | 1.000 |
|  | 18-25 years vs. 45-64 years | -1.0 (-4.1 to 2.1) | 0.524 | 0.918 | 7.0 (4.3 to 9.6) | 0.000 | 0.000 | 21.3 (18.7 to 24.0) | 0.000 | 0.000 | 4.5 (1.7 to 7.2) | 0.001 | 0.004 | -3.0 (-5.9 to -0.0) | 0.049 | 0.148 | 6.6 (3.7 to 9.6) | 0.000 | 0.000 | 3.8 (0.7 to 6.9) | 0.017 | 0.068 | 0.5 (-2.6 to 3.7) | 0.742 | 1.000 | 1.2 (-1.6 to 4.0) | 0.394 | 1.000 |
|  | 18-25 years vs. ≥65 years | 1.1 (-1.9 to 4.1) | 0.459 | 0.918 | 9.9 (7.4 to 12.5) | 0.000 | 0.000 | 29.2 (26.5 to 31.8) | 0.000 | 0.000 | -0.8 (-3.7 to 2.0) | 0.577 | 1.000 | -7.5 (-10.6 to -4.5) | 0.000 | 0.000 | 3.9 (1.1 to 6.7) | 0.007 | 0.020 | 1.7 (-1.2 to 4.7) | 0.250 | 0.250 | -5.8 (-9.1 to -2.6) | 0.000 | 0.002 | 0.1 (-2.7 to 2.8) | 0.957 | 1.000 |
|  | 26-44 years vs. 45-64 years | 1.8 (-0.1 to 3.7) | 0.063 | 0.254 | 4.0 (2.0 to 5.9) | 0.000 | 0.000 | 11.7 (9.7 to 13.7) | 0.000 | 0.000 | -0.5 (-2.5 to 1.5) | 0.623 | 1.000 | -1.9 (-3.9 to 0.1) | 0.057 | 0.148 | 2.3 (0.3 to 4.3) | 0.023 | 0.046 | -2.3 (-4.3 to -0.3) | 0.026 | 0.077 | -0.6 (-2.4 to 1.2) | 0.512 | 1.000 | 0.7 (-1.1 to 2.4) | 0.476 | 1.000 |
|  | 26-44 years vs. ≥65 years | 4.0 (2.1 to 5.8) | 0.000 | 0.000 | 6.9 (5.1 to 8.7) | 0.000 | 0.000 | 19.5 (17.7 to 21.4) | 0.000 | 0.000 | -5.8 (-7.6 to -4.0) | 0.000 | 0.000 | -6.5 (-8.4 to -4.7) | 0.000 | 0.000 | -0.4 (-2.2 to 1.4) | 0.651 | 0.651 | -4.3 (-6.2 to -2.5) | 0.000 | 0.000 | -7.0 (-8.8 to -5.2) | 0.000 | 0.000 | -0.5 (-2.2 to 1.2) | 0.577 | 1.000 |
|  | 45-64 years vs. ≥65 years | 2.1 (0.3 to 3.9) | 0.020 | 0.102 | 2.9 (1.1 to 4.7) | 0.001 | 0.003 | 7.8 (6.0 to 9.7) | 0.000 | 0.000 | -5.3 (-7.1 to -3.5) | 0.000 | 0.000 | -4.6 (-6.4 to -2.8) | 0.000 | 0.000 | -2.7 (-4.6 to -0.8) | 0.005 | 0.020 | -2.0 (-3.9 to -0.1) | 0.036 | 0.077 | -6.4 (-8.1 to -4.6) | 0.000 | 0.000 | -1.1 (-2.8 to 0.5) | 0.179 | 1.000 |
| Race and ethnicity |  |  |  |  |  |  |  |  |  |  |  |  |  |  |  |  |  |  |  |  |  |  |  |  |  |  |  |  |
|  | Hispanic/Latino vs. NH-American Indian/Alaskan Native | 5.2 (-6.0 to 16.5) | 0.361 | 1.000 | 1.4 (-8.8 to 11.6) | 0.786 | 1.000 | 3.1 (-7.5 to 13.7) | 0.565 | 1.000 | 1.8 (-9.4 to 13.0) | 0.755 | 1.000 | 0.2 (-10.8 to 11.2) | 0.971 | 1.000 | 13.0 (3.1 to 22.9) | 0.010 | 0.084 | 2.7 (-6.6 to 11.9) | 0.572 | 1.000 | 7.5 (-2.7 to 17.6) | 0.150 | 0.600 | 12.1 (2.7 to 21.5) | 0.011 | 0.114 |
|  | Hispanic/Latino vs. NH-Asian | 0.2 (-3.5 to 3.9) | 0.919 | 1.000 | -0.5 (-4.3 to 3.3) | 0.808 | 1.000 | -9.3 (-13.4 to -5.2) | 0.000 | 0.000 | -3.1 (-7.0 to 0.8) | 0.120 | 1.000 | -5.5 (-9.3 to -1.8) | 0.004 | 0.051 | -1.7 (-5.1 to 1.8) | 0.352 | 1.000 | -2.5 (-6.6 to 1.5) | 0.224 | 1.000 | -14.7 (-18.4 to -11.0) | 0.000 | 0.000 | -3.0 (-6.1 to 0.1) | 0.059 | 0.415 |
|  | Hispanic/Latino vs. NH-Black/African American | 6.3 (3.0 to 9.6) | 0.000 | 0.003 | 6.2 (3.0 to 9.3) | 0.000 | 0.002 | 3.8 (0.4 to 7.1) | 0.027 | 0.257 | 4.6 (1.5 to 7.7) | 0.003 | 0.042 | 1.7 (-1.7 to 5.1) | 0.314 | 1.000 | 5.0 (1.9 to 8.1) | 0.002 | 0.017 | 3.3 (0.1 to 6.5) | 0.041 | 0.493 | 6.6 (3.4 to 9.7) | 0.000 | 0.000 | 6.9 (4.0 to 9.8) | 0.000 | 0.000 |
|  | Hispanic/Latino vs. NH-Multiple or other race groups | -2.3 (-8.0 to 3.4) | 0.433 | 1.000 | -0.1 (-5.3 to 5.2) | 0.981 | 1.000 | -1.6 (-7.1 to 3.9) | 0.567 | 1.000 | 0.7 (-4.8 to 6.3) | 0.793 | 1.000 | 0.6 (-5.1 to 6.3) | 0.844 | 1.000 | 10.4 (4.9 to 15.9) | 0.000 | 0.003 | 1.2 (-4.7 to 7.1) | 0.688 | 1.000 | 1.4 (-4.9 to 7.6) | 0.667 | 1.000 | 3.1 (-2.3 to 8.4) | 0.258 | 0.773 |
|  | Hispanic/Latino vs. NH-White | -2.6 (-5.1 to -0.1) | 0.042 | 0.459 | -5.9 (-8.1 to -3.7) | 0.000 | 0.000 | -6.8 (-9.2 to -4.4) | 0.000 | 0.000 | -3.5 (-5.8 to -1.3) | 0.002 | 0.025 | -2.2 (-4.5 to 0.1) | 0.061 | 0.614 | 4.6 (2.3 to 6.9) | 0.000 | 0.001 | -2.5 (-4.9 to -0.2) | 0.035 | 0.455 | -5.3 (-7.7 to -3.0) | 0.000 | 0.000 | -1.9 (-4.1 to 0.2) | 0.077 | 0.463 |
|  | NH-American Indian/Alaskan Native vs. NH-Asian | -5.0 (-16.5 to 6.5) | 0.389 | 1.000 | -1.9 (-12.3 to 8.6) | 0.723 | 1.000 | -12.4 (-23.2 to -1.5) | 0.026 | 0.257 | -4.9 (-16.5 to 6.8) | 0.413 | 1.000 | -5.7 (-17.0 to 5.5) | 0.317 | 1.000 | -14.6 (-24.6 to -4.7) | 0.004 | 0.037 | -5.2 (-14.6 to 4.3) | 0.282 | 1.000 | -22.2 (-32.4 to -11.9) | 0.000 | 0.000 | -15.1 (-24.5 to -5.6) | 0.002 | 0.022 |
|  | NH-American Indian/Alaskan Native vs. NH-Black/African American | 1.1 (-10.4 to 12.5) | 0.852 | 1.000 | 4.7 (-5.5 to 15.0) | 0.363 | 1.000 | 0.7 (-10.0 to 11.4) | 0.900 | 1.000 | 2.8 (-8.8 to 14.4) | 0.637 | 1.000 | 1.5 (-9.6 to 12.7) | 0.786 | 1.000 | -8.0 (-17.9 to 1.8) | 0.111 | 0.446 | 0.7 (-8.7 to 10.1) | 0.886 | 1.000 | -0.9 (-11.3 to 9.5) | 0.863 | 1.000 | -5.2 (-14.5 to 4.2) | 0.277 | 0.773 |
|  | NH-American Indian/Alaskan Native vs. NH-Multiple or other race groups | -7.5 (-20.1 to 5.0) | 0.240 | 1.000 | -1.5 (-12.8 to 9.8) | 0.798 | 1.000 | -4.7 (-16.4 to 7.0) | 0.431 | 1.000 | -1.0 (-13.2 to 11.1) | 0.867 | 1.000 | 0.4 (-11.8 to 12.5) | 0.952 | 1.000 | -2.6 (-13.1 to 8.0) | 0.632 | 1.000 | -1.5 (-11.9 to 9.0) | 0.785 | 1.000 | -6.1 (-17.9 to 5.7) | 0.311 | 0.933 | -9.0 (-19.2 to 1.2) | 0.082 | 0.463 |
|  | NH-American Indian/Alaskan Native vs. NH-White | -7.8 (-19.0 to 3.4) | 0.173 | 1.000 | -7.3 (-17.3 to 2.7) | 0.154 | 1.000 | -9.9 (-20.4 to 0.6) | 0.065 | 0.424 | -5.3 (-16.6 to 6.0) | 0.355 | 1.000 | -2.4 (-13.4 to 8.6) | 0.666 | 1.000 | -8.4 (-18.0 to 1.3) | 0.089 | 0.446 | -5.2 (-14.2 to 3.8) | 0.255 | 1.000 | -12.8 (-22.8 to -2.8) | 0.013 | 0.088 | -14.0 (-23.2 to -4.8) | 0.003 | 0.031 |
|  | NH-Asian vs. NH-Black/African American | 6.1 (2.1 to 10.2) | 0.003 | 0.040 | 6.6 (2.7 to 10.6) | 0.001 | 0.012 | 13.1 (9.1 to 17.0) | 0.000 | 0.000 | 7.7 (3.6 to 11.8) | 0.000 | 0.004 | 7.3 (3.2 to 11.4) | 0.001 | 0.008 | 6.6 (3.0 to 10.3) | 0.000 | 0.004 | 5.8 (1.8 to 9.9) | 0.005 | 0.070 | 21.2 (17.5 to 25.0) | 0.000 | 0.000 | 9.9 (6.5 to 13.3) | 0.000 | 0.000 |
|  | NH-Asian vs. NH-Multiple or other race groups | -2.5 (-8.6 to 3.6) | 0.426 | 1.000 | 0.4 (-5.5 to 6.3) | 0.893 | 1.000 | 7.7 (2.0 to 13.3) | 0.008 | 0.083 | 3.8 (-2.7 to 10.4) | 0.252 | 1.000 | 6.1 (0.1 to 12.1) | 0.046 | 0.547 | 12.1 (6.3 to 17.8) | 0.000 | 0.001 | 3.7 (-2.9 to 10.3) | 0.268 | 1.000 | 16.1 (9.4 to 22.7) | 0.000 | 0.000 | 6.0 (0.5 to 11.6) | 0.034 | 0.306 |
|  | NH-Asian vs. NH-White | -2.8 (-6.0 to 0.5) | 0.095 | 0.952 | -5.4 (-8.6 to -2.2) | 0.001 | 0.013 | 2.5 (-0.9 to 5.9) | 0.151 | 0.757 | -0.5 (-4.0 to 3.0) | 0.793 | 1.000 | 3.3 (0.0 to 6.6) | 0.047 | 0.547 | 6.3 (3.1 to 9.4) | 0.000 | 0.001 | -0.0 (-3.5 to 3.4) | 0.982 | 1.000 | 9.4 (6.1 to 12.7) | 0.000 | 0.000 | 1.0 (-1.6 to 3.7) | 0.439 | 0.773 |
|  | NH-Black/African American vs. NH-Multiple or other race groups | -8.6 (-14.4 to -2.8) | 0.004 | 0.043 | -6.2 (-11.6 to -0.8) | 0.025 | 0.234 | -5.4 (-11.0 to 0.2) | 0.061 | 0.424 | -3.8 (-9.7 to 2.0) | 0.200 | 1.000 | -1.2 (-7.2 to 4.9) | 0.702 | 1.000 | 5.4 (-0.2 to 11.1) | 0.059 | 0.351 | -2.1 (-8.3 to 4.0) | 0.495 | 1.000 | -5.2 (-11.4 to 1.1) | 0.104 | 0.518 | -3.8 (-9.1 to 1.4) | 0.153 | 0.614 |
|  | NH-Black/African American vs. NH-White | -8.9 (-11.6 to -6.2) | 0.000 | 0.000 | -12.0 (-14.7 to -9.4) | 0.000 | 0.000 | -10.6 (-13.3 to -7.8) | 0.000 | 0.000 | -8.1 (-10.7 to -5.5) | 0.000 | 0.000 | -4.0 (-6.6 to -1.3) | 0.004 | 0.051 | -0.3 (-2.9 to 2.2) | 0.793 | 1.000 | -5.9 (-8.5 to -3.3) | 0.000 | 0.000 | -11.9 (-14.4 to -9.4) | 0.000 | 0.000 | -8.9 (-11.3 to -6.4) | 0.000 | 0.000 |
|  | NH-Multiple or other race groups vs. NH-White | -0.3 (-5.6 to 5.0) | 0.914 | 1.000 | -5.8 (-10.8 to -0.8) | 0.023 | 0.234 | -5.2 (-10.2 to -0.1) | 0.044 | 0.352 | -4.3 (-9.8 to 1.2) | 0.127 | 1.000 | -2.8 (-8.1 to 2.6) | 0.306 | 1.000 | -5.8 (-10.9 to -0.7) | 0.027 | 0.190 | -3.7 (-9.3 to 1.8) | 0.187 | 1.000 | -6.7 (-12.5 to -0.8) | 0.025 | 0.151 | -5.0 (-10.1 to 0.0) | 0.052 | 0.415 |
| Abbreviations: CI, Confidence Interval; NH, non-Hispanic; WPHI, Whole Person Health Index | | | | | | | | | | | | | | | | | | | | | | | | | | | | |
| Note: Results show the survey-weighted contrasts of age-standardized (except for age groups) proportions with Holm adjustment for multiple comparisons. Comparisons are of individuals responding 'excellent' or 'very good' (vs. 'good'/'fair'/'poor') within each group. | | | | | | | | | | | | | | | | | | | | | | | | | | | | |
| ^a^ Participants responded to the following questions that were asked in the following order, using a 5-point Likert scale of 1 (excellent), 2 (very good), 3 (good), 4 (fair), or 5 (poor): Would you say your health in general is...? How would you rate your quality of life, focusing on what matters most to you? How would you rate your social and family connections? In general, how healthy is your overall diet? How would you rate your physical activity, compared with people in your age group? How would you rate your ability to manage stress? How would you rate your sleep? How would you rate your ability to find meaning and purpose in your daily life? How would you rate your ability to manage your health, focusing on aspects of your health that matter most to you?. Responses were reverse coded so that higher scores indicated higher levels of health. To estimate average whole person health, the nine individual items were averaged, rounded to the nearest integer, and dichotomized as ≥4 ('excellent' or 'very good') vs. ≤3 ('good', 'fair', or 'poor'). | | | | | | | | | | | | | | | | | | | | | | | | | | | | |

### Supplemental Table 3. Prevalence of “optimal” (defined as ‘excellent’, ‘very good’, or ‘good’) whole person health and whole person health components by sociodemographic characteristics, National Health Interview Survey, 2025 (N=23,884)

| Sociodemographic characteristic | Unweighted n | Prevalence (95% CI) of an average response of 'excellent', 'very good', or 'good' to the Whole Person Health Index ^a, b^ | Prevalence (95% CI) of endorsement of 'excellent', 'very good', or ‘good’ to questions/components of the Whole Person Health Index (WPHI) ^a^ | | | | | | | | |
| --- | --- | --- | --- | --- | --- | --- | --- | --- | --- | --- | --- |
|  |  |  | Meaning/  Purpose ^b,c^ | Quality of Life ^b, c, d^ | General Health ^b, d^ | Management of Health ^b, d^ | Stress Management ^b, c, d^ | Sleep ^b, c, d^ | Physical Activity ^b,c^ | Diet ^b, c, d^ | Social/Family Connections ^b,c^ |
| Overall | 23,884 | 92.7 (92.3-93.2) | 91.1  (90.6-91.6) | 92.1  (91.6-92.5) | 85.5  (84.9-86.1) | 89.3  (88.8-89.8) | 84.7  (84.2-85.3) | 71.7  (70.9-72.4) | 78.6  (78.0-79.3) | 84.0  (83.3-84.7) | 92.0  (91.5-92.5) |
| Age group |  |  |  |  |  |  |  |  |  |  |  |
| 18-25 years | 1,634 | 93.8 (91.6-96.0) | 88.9  (85.7-92.0) | 92.0  (89.0-94.9) | 94.2  (92.1-96.3) | 86.1  (82.8-89.5) | 78.2  (74.2-82.1) | 75.0  (71.0-79.0) | 78.3  (74.4-82.2) | 80.8  (77.2-84.4) | 91.3  (88.5-94.0) |
| 26-44 years | 6,788 | 93.0 (92.2-93.8) | 91.1  (90.3-91.9) | 93.1  (92.4-93.9) | 90.4  (89.5-91.3) | 87.9  (87.0-88.9) | 82.6  (81.5-83.7) | 71.3  (70.0-72.6) | 77.7  (76.5-78.9) | 82.5  (81.4-83.6) | 91.6  (90.8-92.5) |
| 45-64 years | 7,125 | 92.1 (91.3-92.9) | 91.6  (90.9-92.4) | 91.3  (90.5-92.1) | 83.0  (81.8-84.1) | 89.5  (88.7-90.3) | 85.6  (84.7-86.6) | 69.1  (67.8-70.4) | 79.3  (78.1-80.4) | 83.6  (82.5-84.8) | 92.0  (91.3-92.8) |
| ≥ 65 years | 8,337 | 92.7 (92.0-93.4) | 91.3  (90.6-92.0) | 90.5  (89.7-91.3) | 77.0  (75.8-78.1) | 91.6  (90.9-92.3) | 89.3  (88.4-90.1) | 73.9  (72.7-75.0) | 78.1  (77.1-79.1) | 88.7  (87.9-89.5) | 93.0  (92.3-93.6) |
| Sex |  |  |  |  |  |  |  |  |  |  |  |
| Men | 11,071 | 92.7 (92.1-93.3) | 90.0  (89.3-90.7) | 91.4  (90.8-92.1) | 85.5  (84.8-86.3) | 88.8  (88.1-89.6) | 86.3  (85.5-87.1) | 73.1  (72.0-74.1) | 81.0  (80.1-81.9) | 83.0  (82.1-84.0) | 91.2  (90.5-91.9) |
| Women | 12,813 | 92.7 (92.2-93.3) | 92.2  (91.6-92.8) | 92.7  (92.1-93.3) | 85.4  (84.6-86.2) | 89.7  (89.0-90.4) | 83.2  (82.4-84.0) | 70.4  (69.3-71.4) | 76.4  (75.4-77.3) | 85.0  (84.2-85.9) | 92.8  (92.2-93.3) |
| Race and ethnicity |  |  |  |  |  |  |  |  |  |  |  |
| Hispanic/Latino | 3,364 | 92.4 (91.2-93.5) | 91.7  (90.5-93.0) | 90.9  (89.7-92.1) | 82.0  (80.3-83.8) | 88.6  (87.3-89.8) | 84.7  (83.3-86.1) | 73.3  (71.6-75.1) | 77.5  (75.7-79.2) | 81.5  (79.9-83.0) | 93.0  (91.9-94.1) |
| NH-American Indian/Alaskan Native | 158 | 89.5 (83.6-95.4) | 88.1  (81.5-94.7) | 86.0  (78.3-93.7) | 73.5  (64.9-82.1) | 91.1  (86.3-95.9) | 76.9  (68.5-85.3) | 62.7  (54.1-71.3) | 73.7  (65.4-82.0) | 82.9  (76.0-89.8) | 92.0  (88.4-95.7) |
| NH-Asian | 1,431 | 95.4 (94.2-96.6) | 91.4  (89.8-93.1) | 93.7  (92.1-95.2) | 87.1  (84.9-89.3) | 91.7  (90.1-93.2) | 90.4  (88.7-92.0) | 76.1  (73.6-78.6) | 83.6  (81.2-85.9) | 91.0  (89.1-92.9) | 94.6  (93.3-95.9) |
| NH-Black/African American | 2,680 | 90.2 (88.9-91.6) | 89.0  (87.5-90.4) | 88.5  (87.1-90.0) | 80.4  (78.5-82.2) | 87.2  (85.6-88.7) | 81.5  (79.7-83.3) | 68.8  (66.7-70.8) | 74.9  (72.9-76.9) | 78.7  (76.7-80.7) | 88.5  (87.1-90.0) |
| NH-Multiple or other race groups | 455 | 87.7 (84.1-91.4) | 89.7  (86.5-92.9) | 85.9  (82.0-89.7) | 83.5  (79.9-87.2) | 86.3  (82.7-90.0) | 82.9  (78.8-87.0) | 61.3  (55.8-66.8) | 73.5  (68.5-78.4) | 81.6  (77.0-86.3) | 88.5  (85.1-91.8) |
| NH-White | 15,796 | 93.0 (92.5-93.6) | 91.3  (90.7-91.8) | 93.0  (92.5-93.5) | 87.1  (86.4-87.8) | 89.5  (88.9-90.2) | 84.8  (84.0-85.6) | 71.4  (70.4-72.4) | 79.4  (78.5-80.3) | 85.1  (84.3-86.0) | 92.2  (91.6-92.7) |
| Abbreviations: CI, Confidence Interval; NH, non-Hispanic; WPHI, Whole Person Health Index | | | | | | | | | | | |
| Note: Cells contain the survey-weighted, age-standardized to the 2020 Census prevalence or proportion (95% confidence interval) of individuals responding 'excellent', 'very good', or ‘good’ (vs. 'fair'/'poor'). | | | | | | | | | | | |
| ^a^ Participants responded to the following questions that were asked in the following order, using a 5-point Likert scale of 1 (excellent), 2 (very good), 3 (good), 4 (fair), or 5 (poor): Would you say your health in general is…? How would you rate your quality of life, focusing on what matters most to you? How would you rate your social and family connections? In general, how healthy is your overall diet? How would you rate your physical activity, compared with people in your age group? How would you rate your ability to manage stress? How would you rate your sleep? How would you rate your ability to find meaning and purpose in your daily life? How would you rate your ability to manage your health, focusing on aspects of your health that matter most to you?. Responses were reverse coded so that higher scores indicated higher levels of health; scores were then dichotomized as ≥4 ('excellent' or 'very good') vs. '≤3 'good', 'fair', or 'poor'. To estimate average whole person health, the nine individual items were averaged, rounded to the nearest integer and dichotomized as ≥3 ('excellent', 'very good', or 'good') vs. ≤2 ('fair' or 'poor'. | | | | | | | | | | | |
| ^b^ p-value for the omnibus Rao-Scott p-value <0.05, indicating differences between race and ethnicity groups. | | | | | | | | | | | |
| ^c^ p-value for the omnibus Rao-Scott p-value <0.05, indicating differences between sex. | | | | | | | | | | | |
| ^d^ p-value for the omnibus Rao-Scott p-value <0.05, indicating differences between age groups. | | | | | | | | | | | |

### Supplemental Table 4. Pairwise comparisons of the prevalence of “optimal” (defined as ‘excellent’, ‘very good’, or ‘good’) whole person health by sociodemographic characteristics, National Health Interview Survey, 2025 (N=23,884)

| Sociodemographic characteristic | Pairwise comparison | Prevalence difference (95% CI) of an average response of 'excellent', 'very good', or 'good' to the Whole Person Health Index ^a^ | p-value | Holm adjusted p-value |
| --- | --- | --- | --- | --- |
| Age group |  |  |  |  |
|  | 18-25 years vs. 26-44 years | 0.6 (-0.9 to 2.2) | 0.419 | 0.875 |
|  | 18-25 years vs. 45-64 years | 1.5 (0.0 to 3.1) | 0.048 | 0.287 |
|  | 18-25 years vs. ≥65 years | 0.9 (-0.6 to 2.5) | 0.219 | 0.875 |
|  | 26-44 years vs. 45-64 years | 0.9 (-0.2 to 2.0) | 0.099 | 0.495 |
|  | 26-44 years vs. ≥65 years | 0.3 (-0.7 to 1.3) | 0.539 | 0.875 |
|  | 45-64 years vs. ≥65 years | -0.6 (-1.6 to 0.4) | 0.261 | 0.875 |
| Race and ethnicity |  |  |  |  |
|  | Hispanic/Latino vs. NH-American Indian/Alaskan Native | 1.9 (-3.4 to 7.2) | 0.481 | 1.000 |
|  | Hispanic/Latino vs. NH-Asian | -2.8 (-4.4 to -1.1) | 0.001 | 0.013 |
|  | Hispanic/Latino vs. NH-Black/African American | 2.3 (0.5 to 4.0) | 0.013 | 0.131 |
|  | Hispanic/Latino vs. NH-Multiple or other race groups | 3.8 (0.3 to 7.2) | 0.034 | 0.271 |
|  | Hispanic/Latino vs. NH-White | -0.4 (-1.7 to 0.8) | 0.474 | 1.000 |
|  | NH-American Indian/Alaskan Native vs. NH-Asian | -4.7 (-10.0 to 0.7) | 0.085 | 0.596 |
|  | NH-American Indian/Alaskan Native vs. NH-Black/African American | 0.3 (-5.0 to 5.7) | 0.898 | 1.000 |
|  | NH-American Indian/Alaskan Native vs. NH-Multiple or other race groups | 1.8 (-4.5 to 8.2) | 0.568 | 1.000 |
|  | NH-American Indian/Alaskan Native vs. NH-White | -2.4 (-7.6 to 2.8) | 0.373 | 1.000 |
|  | NH-Asian vs. NH-Black/African American | 5.0 (3.2 to 6.9) | 0.000 | 0.000 |
|  | NH-Asian vs. NH-Multiple or other race groups | 6.5 (2.8 to 10.2) | 0.001 | 0.007 |
|  | NH-Asian vs. NH-White | 2.3 (1.0 to 3.6) | 0.001 | 0.007 |
|  | NH-Black/African American vs. NH-Multiple or other race groups | 1.5 (-2.1 to 5.1) | 0.421 | 1.000 |
|  | NH-Black/African American vs. NH-White | -2.7 (-4.2 to -1.2) | 0.000 | 0.004 |
|  | NH-Multiple or other race groups vs. NH-White | -4.2 (-7.7 to -0.7) | 0.017 | 0.157 |
| Abbreviations: CI, Confidence Interval; NH, non-Hispanic; WPHI, Whole Person Health Index | | | | |
| Note: Results show the survey-weighted contrasts of age-standardized (except for age groups) proportions with Holm adjustment for multiple comparisons. Comparisons are of individuals responding 'excellent', 'very good', or 'good' (vs. 'fair'/'poor') within each group. | | | | |
| ^a^ Participants responded to the following questions that were asked in the following order, using a 5-point Likert scale of 1 (excellent), 2 (very good), 3 (good), 4 (fair), or 5 (poor): Would you say your health in general is…? How would you rate your quality of life, focusing on what matters most to you? How would you rate your social and family connections? In general, how healthy is your overall diet? How would you rate your physical activity, compared with people in your age group? How would you rate your ability to manage stress? How would you rate your sleep? How would you rate your ability to find meaning and purpose in your daily life? How would you rate your ability to manage your health, focusing on aspects of your health that matter most to you?. Responses were reverse coded so that higher scores indicated higher levels of health. To estimate average whole person health, the nine individual items were averaged, rounded to the nearest integer and dichotomized as ≥3 ('excellent', 'very good', or 'good') vs. '≤2 ('fair' or 'poor'). | | | | |

### Supplemental Table 5. Pairwise comparisons of the prevalence of “optimal” (defined as ‘excellent’, ‘very good’, or ‘good’) whole person health components by sociodemographic characteristics, National Health Interview Survey, 2025 (N=23884)

| Sociodemographic characteristic | Pairwise comparison | Endorsement of 'excellent', 'very good', or 'good' to questions/components of the Whole Person Health Index (WPHI) ^a^ | | | | | | | | | | | | | | | | | | | | | | | | | | |
| --- | --- | --- | --- | --- | --- | --- | --- | --- | --- | --- | --- | --- | --- | --- | --- | --- | --- | --- | --- | --- | --- | --- | --- | --- | --- | --- | --- | --- |
|  |  | Meaning/Purpose | | | Quality of Life | | | General Health | | | Management of Health | | | Stress Management | | | Sleep | | | Physical Activity | | | Diet | | | Social/Family Connections | | |
|  |  | Prevalence difference (95% CI) | p-value | Holm adjusted p-value | Prevalence difference (95% CI) | p-value | Holm adjusted p-value | Prevalence difference (95% CI) | p-value | Holm adjusted p-value | Prevalence difference (95% CI) | p-value | Holm adjusted p-value | Prevalence difference (95% CI) | p-value | Holm adjusted p-value | Prevalence difference (95% CI) | p-value | Holm adjusted p-value | Prevalence difference (95% CI) | p-value | Holm adjusted p-value | Prevalence difference (95% CI) | p-value | Holm adjusted p-value | Prevalence difference (95% CI) | p-value | Holm adjusted p-value |
| Age group |  |  |  |  |  |  |  |  |  |  |  |  |  |  |  |  |  |  |  |  |  |  |  |  |  |  |  |  |
|  | 18-25 years vs. 26-44 years | -1.6 (-3.5 to 0.2) | 0.087 | 0.348 | 0.9 (-0.6 to 2.4) | 0.250 | 0.320 | 3.6 (2.1 to 5.2) | 0.000 | 0.000 | 0.5 (-1.4 to 2.4) | 0.619 | 0.619 | -2.4 (-4.9 to 0.1) | 0.062 | 0.062 | 3.7 (1.1 to 6.3) | 0.006 | 0.017 | 2.5 (0.1 to 4.9) | 0.039 | 0.232 | -1.4 (-3.9 to 1.0) | 0.242 | 0.265 | -0.3 (-2.1 to 1.5) | 0.750 | 1.000 |
|  | 18-25 years vs. 45-64 years | -2.1 (-4.0 to -0.2) | 0.027 | 0.165 | 2.7 (1.2 to 4.3) | 0.001 | 0.002 | 11.0 (9.4 to 12.7) | 0.000 | 0.000 | -1.1 (-2.9 to 0.8) | 0.250 | 0.501 | -5.4 (-7.8 to -3.0) | 0.000 | 0.000 | 5.9 (3.3 to 8.5) | 0.000 | 0.000 | 1.0 (-1.4 to 3.4) | 0.419 | 0.838 | -2.6 (-5.0 to -0.1) | 0.040 | 0.119 | -0.7 (-2.4 to 1.0) | 0.426 | 1.000 |
|  | 18-25 years vs. ≥65 years | -1.8 (-3.6 to -0.0) | 0.046 | 0.228 | 3.5 (2.0 to 5.0) | 0.000 | 0.000 | 17.0 (15.3 to 18.7) | 0.000 | 0.000 | -3.2 (-5.0 to -1.4) | 0.001 | 0.002 | -9.0 (-11.3 to -6.6) | 0.000 | 0.000 | 1.2 (-1.4 to 3.7) | 0.371 | 0.371 | 2.1 (-0.3 to 4.6) | 0.082 | 0.331 | -7.7 (-10.0 to -5.4) | 0.000 | 0.000 | -1.6 (-3.4 to 0.1) | 0.068 | 0.272 |
|  | 26-44 years vs. 45-64 years | -0.5 (-1.6 to 0.6) | 0.386 | 1.000 | 1.9 (0.8 to 2.9) | 0.000 | 0.002 | 7.4 (6.1 to 8.8) | 0.000 | 0.000 | -1.6 (-2.8 to -0.4) | 0.011 | 0.032 | -3.0 (-4.5 to -1.5) | 0.000 | 0.000 | 2.2 (0.5 to 3.9) | 0.010 | 0.019 | -1.6 (-3.2 to 0.1) | 0.066 | 0.331 | -1.1 (-2.6 to 0.3) | 0.133 | 0.265 | -0.4 (-1.5 to 0.7) | 0.494 | 1.000 |
|  | 26-44 years vs. ≥65 years | -0.2 (-1.2 to 0.9) | 0.721 | 1.000 | 2.6 (1.6 to 3.7) | 0.000 | 0.000 | 13.4 (12.0 to 14.7) | 0.000 | 0.000 | -3.7 (-4.8 to -2.5) | 0.000 | 0.000 | -6.6 (-7.9 to -5.3) | 0.000 | 0.000 | -2.5 (-4.2 to -0.9) | 0.003 | 0.012 | -0.4 (-1.9 to 1.2) | 0.625 | 0.838 | -6.3 (-7.6 to -4.9) | 0.000 | 0.000 | -1.3 (-2.4 to -0.3) | 0.013 | 0.078 |
|  | 45-64 years vs. ≥65 years | 0.3 (-0.7 to 1.3) | 0.555 | 1.000 | 0.8 (-0.3 to 1.9) | 0.160 | 0.320 | 6.0 (4.6 to 7.4) | 0.000 | 0.000 | -2.1 (-3.2 to -1.0) | 0.000 | 0.001 | -3.6 (-4.9 to -2.4) | 0.000 | 0.000 | -4.7 (-6.5 to -3.0) | 0.000 | 0.000 | 1.2 (-0.3 to 2.6) | 0.123 | 0.370 | -5.1 (-6.5 to -3.8) | 0.000 | 0.000 | -0.9 (-1.9 to -0.0) | 0.049 | 0.247 |
| Race and ethnicity |  |  |  |  |  |  |  |  |  |  |  |  |  |  |  |  |  |  |  |  |  |  |  |  |  |  |  |  |
|  | Hispanic/Latino vs. NH-American Indian/Alaskan Native | 3.6 (-3.8 to 10.9) | 0.342 | 1.000 | 4.1 (-3.1 to 11.4) | 0.262 | 1.000 | 8.6 (0.7 to 16.5) | 0.032 | 0.194 | -3.1 (-7.8 to 1.7) | 0.208 | 1.000 | 6.5 (-2.2 to 15.1) | 0.141 | 0.889 | 10.9 (1.6 to 20.3) | 0.021 | 0.128 | 2.4 (-6.0 to 10.7) | 0.575 | 1.000 | -2.0 (-9.5 to 5.5) | 0.600 | 1.000 | 0.3 (-3.1 to 3.6) | 0.869 | 1.000 |
|  | Hispanic/Latino vs. NH-Asian | 0.3 (-1.7 to 2.2) | 0.785 | 1.000 | -2.0 (-3.9 to -0.2) | 0.030 | 0.271 | -3.6 (-6.4 to -0.9) | 0.009 | 0.086 | -2.9 (-5.0 to -0.7) | 0.008 | 0.110 | -6.1 (-8.2 to -4.0) | 0.000 | 0.000 | -2.7 (-5.7 to 0.3) | 0.074 | 0.233 | -5.6 (-8.6 to -2.6) | 0.000 | 0.004 | -9.8 (-12.2 to -7.3) | 0.000 | 0.000 | -1.5 (-3.2 to 0.2) | 0.075 | 0.603 |
|  | Hispanic/Latino vs. NH-Black/African American | 2.8 (0.8 to 4.7) | 0.005 | 0.069 | 3.0 (1.2 to 4.8) | 0.001 | 0.015 | 3.1 (0.7 to 5.4) | 0.012 | 0.100 | 1.4 (-0.7 to 3.4) | 0.186 | 1.000 | 2.7 (0.3 to 5.0) | 0.026 | 0.235 | 4.8 (2.1 to 7.5) | 0.000 | 0.005 | 2.5 (-0.2 to 5.1) | 0.065 | 0.583 | 2.5 (-0.0 to 5.1) | 0.050 | 0.418 | 4.6 (2.8 to 6.4) | 0.000 | 0.000 |
|  | Hispanic/Latino vs. NH-Multiple or other race groups | 2.2 (-1.2 to 5.6) | 0.210 | 1.000 | 4.8 (1.1 to 8.5) | 0.011 | 0.114 | -2.3 (-6.0 to 1.3) | 0.204 | 0.817 | 2.5 (-1.5 to 6.4) | 0.223 | 1.000 | 2.2 (-2.3 to 6.7) | 0.330 | 1.000 | 12.5 (6.4 to 18.5) | 0.000 | 0.001 | 4.3 (-0.9 to 9.4) | 0.102 | 0.614 | 0.1 (-5.2 to 5.4) | 0.973 | 1.000 | 3.5 (0.2 to 6.9) | 0.037 | 0.336 |
|  | Hispanic/Latino vs. NH-White | 0.5 (-0.8 to 1.8) | 0.475 | 1.000 | -1.1 (-2.3 to 0.0) | 0.057 | 0.456 | -2.2 (-3.8 to -0.5) | 0.013 | 0.100 | -1.3 (-2.7 to 0.1) | 0.079 | 0.627 | -1.2 (-2.8 to 0.4) | 0.135 | 0.889 | 2.1 (0.2 to 4.0) | 0.035 | 0.173 | -1.7 (-3.6 to 0.2) | 0.074 | 0.583 | -4.5 (-6.2 to -2.8) | 0.000 | 0.000 | 0.7 (-0.5 to 1.9) | 0.254 | 1.000 |
|  | NH-American Indian/Alaskan Native vs. NH-Asian | -3.3 (-10.6 to 4.1) | 0.382 | 1.000 | -6.2 (-13.4 to 1.1) | 0.096 | 0.670 | -12.3 (-20.1 to -4.4) | 0.002 | 0.030 | 0.2 (-4.7 to 5.1) | 0.935 | 1.000 | -12.6 (-21.2 to -4.0) | 0.004 | 0.043 | -13.7 (-23.0 to -4.3) | 0.004 | 0.038 | -8.0 (-16.5 to 0.6) | 0.067 | 0.583 | -7.7 (-15.4 to -0.1) | 0.046 | 0.418 | -1.8 (-5.3 to 1.7) | 0.310 | 1.000 |
|  | NH-American Indian/Alaskan Native vs. NH-Black/African American | -0.8 (-8.2 to 6.6) | 0.835 | 1.000 | -1.1 (-8.3 to 6.1) | 0.761 | 1.000 | -5.6 (-13.4 to 2.2) | 0.161 | 0.807 | 4.4 (-0.5 to 9.4) | 0.078 | 0.627 | -3.8 (-12.5 to 4.9) | 0.390 | 1.000 | -6.1 (-15.6 to 3.3) | 0.201 | 0.401 | 0.1 (-8.3 to 8.5) | 0.984 | 1.000 | 4.6 (-3.1 to 12.2) | 0.243 | 1.000 | 4.3 (0.8 to 7.8) | 0.016 | 0.162 |
|  | NH-American Indian/Alaskan Native vs. NH-Multiple or other race groups | -1.4 (-9.3 to 6.5) | 0.734 | 1.000 | 0.6 (-7.3 to 8.6) | 0.874 | 1.000 | -11.0 (-19.4 to -2.5) | 0.011 | 0.100 | 5.5 (-0.3 to 11.3) | 0.061 | 0.613 | -4.3 (-13.9 to 5.4) | 0.388 | 1.000 | 1.5 (-9.3 to 12.4) | 0.784 | 0.784 | 1.9 (-7.5 to 11.3) | 0.690 | 1.000 | 2.1 (-7.0 to 11.1) | 0.649 | 1.000 | 3.3 (-1.4 to 7.9) | 0.169 | 1.000 |
|  | NH-American Indian/Alaskan Native vs. NH-White | -3.1 (-10.3 to 4.1) | 0.402 | 1.000 | -5.3 (-12.4 to 1.9) | 0.147 | 0.883 | -10.8 (-18.4 to -3.1) | 0.006 | 0.070 | 1.8 (-2.9 to 6.5) | 0.452 | 1.000 | -7.7 (-16.2 to 0.8) | 0.077 | 0.615 | -8.9 (-18.1 to 0.3) | 0.058 | 0.233 | -4.1 (-12.4 to 4.1) | 0.329 | 1.000 | -2.5 (-9.9 to 4.9) | 0.510 | 1.000 | 0.4 (-2.8 to 3.6) | 0.808 | 1.000 |
|  | NH-Asian vs. NH-Black/African American | 2.5 (0.3 to 4.7) | 0.023 | 0.298 | 5.1 (3.0 to 7.1) | 0.000 | 0.000 | 6.7 (3.8 to 9.6) | 0.000 | 0.000 | 4.2 (2.0 to 6.5) | 0.000 | 0.003 | 8.8 (6.4 to 11.2) | 0.000 | 0.000 | 7.5 (4.5 to 10.6) | 0.000 | 0.000 | 8.1 (4.9 to 11.3) | 0.000 | 0.000 | 12.3 (9.5 to 15.1) | 0.000 | 0.000 | 6.1 (4.2 to 8.1) | 0.000 | 0.000 |
|  | NH-Asian vs. NH-Multiple or other race groups | 1.9 (-1.9 to 5.7) | 0.322 | 1.000 | 6.8 (2.8 to 10.9) | 0.001 | 0.013 | 1.3 (-2.7 to 5.3) | 0.521 | 1.000 | 5.3 (1.1 to 9.5) | 0.013 | 0.153 | 8.3 (3.7 to 13.0) | 0.001 | 0.006 | 15.2 (8.9 to 21.5) | 0.000 | 0.000 | 9.9 (4.3 to 15.5) | 0.001 | 0.007 | 9.8 (4.3 to 15.4) | 0.001 | 0.005 | 5.1 (1.5 to 8.6) | 0.005 | 0.057 |
|  | NH-Asian vs. NH-White | 0.2 (-1.5 to 1.9) | 0.814 | 1.000 | 0.9 (-0.7 to 2.5) | 0.260 | 1.000 | 1.5 (-0.9 to 3.9) | 0.221 | 0.817 | 1.6 (-0.1 to 3.3) | 0.068 | 0.613 | 4.9 (3.1 to 6.7) | 0.000 | 0.000 | 4.8 (2.2 to 7.4) | 0.000 | 0.004 | 3.9 (1.3 to 6.5) | 0.004 | 0.039 | 5.3 (3.2 to 7.3) | 0.000 | 0.000 | 2.2 (0.8 to 3.6) | 0.002 | 0.030 |
|  | NH-Black/African American vs. NH-Multiple or other race groups | -0.6 (-4.3 to 3.1) | 0.752 | 1.000 | 1.8 (-2.2 to 5.7) | 0.383 | 1.000 | -5.4 (-9.3 to -1.5) | 0.006 | 0.070 | 1.1 (-3.0 to 5.2) | 0.607 | 1.000 | -0.5 (-5.2 to 4.3) | 0.849 | 1.000 | 7.7 (1.5 to 13.9) | 0.016 | 0.125 | 1.8 (-3.5 to 7.2) | 0.505 | 1.000 | -2.5 (-7.8 to 2.9) | 0.373 | 1.000 | -1.1 (-4.5 to 2.4) | 0.543 | 1.000 |
|  | NH-Black/African American vs. NH-White | -2.3 (-3.8 to -0.7) | 0.004 | 0.056 | -4.2 (-5.7 to -2.6) | 0.000 | 0.000 | -5.2 (-7.2 to -3.2) | 0.000 | 0.000 | -2.6 (-4.3 to -1.0) | 0.002 | 0.027 | -3.9 (-5.9 to -2.0) | 0.000 | 0.001 | -2.7 (-5.0 to -0.5) | 0.016 | 0.125 | -4.2 (-6.4 to -2.0) | 0.000 | 0.002 | -7.0 (-9.2 to -4.9) | 0.000 | 0.000 | -3.9 (-5.5 to -2.3) | 0.000 | 0.000 |
|  | NH-Multiple or other race groups vs. NH-White | -1.7 (-5.0 to 1.6) | 0.312 | 1.000 | -5.9 (-9.6 to -2.2) | 0.002 | 0.018 | 0.2 (-3.2 to 3.6) | 0.910 | 1.000 | -3.7 (-7.5 to 0.1) | 0.054 | 0.598 | -3.4 (-7.9 to 1.0) | 0.127 | 0.889 | -10.4 (-16.4 to -4.4) | 0.001 | 0.007 | -6.0 (-11.1 to -0.9) | 0.020 | 0.201 | -4.6 (-9.7 to 0.5) | 0.075 | 0.526 | -2.9 (-6.1 to 0.4) | 0.083 | 0.603 |
| Abbreviations: CI, Confidence Interval; NH, non-Hispanic; WPHI, Whole Person Health Index | | | | | | | | | | | | | | | | | | | | | | | | | | | | |
| Note: Results show the survey-weighted contrasts of age-standardized (except for age groups) proportions with Holm adjustment for multiple comparisons. Comparisons are of individuals responding 'excellent', 'very good', or 'good' (vs. 'fair'/'poor') within each group. | | | | | | | | | | | | | | | | | | | | | | | | | | | | |
| ^a^ Participants responded to the following questions that were asked in the following order, using a 5-point Likert scale of 1 (excellent), 2 (very good), 3 (good), 4 (fair), or 5 (poor): Would you say your health in general is...? How would you rate your quality of life, focusing on what matters most to you? How would you rate your social and family connections? In general, how healthy is your overall diet? How would you rate your physical activity, compared with people in your age group? How would you rate your ability to manage stress? How would you rate your sleep? How would you rate your ability to find meaning and purpose in your daily life? How would you rate your ability to manage your health, focusing on aspects of your health that matter most to you?. Responses were reverse coded so that higher scores indicated higher levels of health. To estimate average whole person health, the nine individual items were averaged, rounded to the nearest integer, and dichotomized as ≥4 ('excellent' or 'very good') vs. ≤3 ('good', 'fair', or 'poor'). | | | | | | | | | | | | | | | | | | | | | | | | | | | | |
